# Harnessing Raman spectroscopy and Multimodal Imaging of Cartilage for Osteoarthritis Diagnosis

**DOI:** 10.1101/2023.09.05.23294936

**Authors:** Anna Crisford, Hiroki Cook, Konstantinos Bourdakos, Seshasailam Venkateswaran, Douglas Dunlop, Richard OC Oreffo, Sumeet Mahajan

## Abstract

Osteoarthritis (OA) is a complex disease of cartilage characterised by chronic joint pain, limitations in mobility and function leading to reduced quality of life. Current methods to diagnose OA, such as X-ray, MRI and the invasive synovial fluid analysis lack molecular specificity and are limited to detection of the late stages of the disease. A rapid minimally invasive and non-destructive approach for early diagnosis of OA is a critical unmet need. Label-free techniques such as Raman Spectroscopy (RS), Coherent anti-Stokes Raman scattering (CARS), Second Harmonic Generation (SHG) and Two Photon Fluorescence (TPF) are increasingly being explored to characterise cartilage tissue. However, current studies are based on whole tissue analysis and do not take into account the different and structurally distinct layers in cartilage. In this work, we used Raman spectroscopy to obtain signatures from superficial and deep layers of healthy and osteoarthritic cartilage obtained from a total of 64 patients (45 OA and 19 controls). Spectra were acquired both in the ‘fingerprint’ region from 700 to 1720 cm^-1^ and high-frequency stretching region from 2500 to 3300 cm^-1^. Principal component and linear discriminant analysis was used to identify the peaks that contributed the most to classification of the different samples. The most pronounced differences were observed at the proline (855 cm^-1^ and 921 cm^-1^) and hydroxyproline (877 cm^-1^ and 938 cm^-1^), sulphated glycosaminoglycan (sGAG) (1064 cm^-1^ and 1380 cm^-1^) for both control and OA as well as the 1245 cm^-1^ and 1272 cm^-1^, 1320 cm^- 1^ and 1345 cm^-1^, 1451 collagen modes in OA samples, consistent with expected collagen structural changes. Classification accuracy based on Raman fingerprint spectral analysis of superficial and deep layer cartilage for controls was found to be 94% and 96%, respectively. OA diseased cartilage was classified with 80% and 87% accuracy based on analysis of the superficial and the deep layers, respectively. Raman spectra from the C-H stretching region (2500-3300 cm^-1^) did not result in high classification accuracies for OA diseased cartilage. Intriguingly, relatively less differences were found with gender in healthy cartilage indicating that OA brings about significant chemical changes across both genders in both layers. On the other hand, we found significant differences in superficial and deep layer cartilage signatures with age (under 60 and over 60 years). Preliminary images of different layers of cartilage using CARS, SHG and TPF showed Cell clustering in OA, and differences in pericellular matrix and collagen structure in the superficial and the deep layers. The current study demonstrates the potential of Raman Spectroscopy together with multimodal imaging as a potential tool that provides insight into the chemical and structural composition of different layers of cartilage to improve OA diagnosis.

## 1. Introduction

Musculoskeletal disorders are the second most common cause of disability worldwide. Osteoarthritis (OA), is the most common form of arthritis, resulting in an estimated 8.75 million of the UK population over 45 years of age seeking treatment annually; and a global prevalence of over 500 million (Hunter, March and Chew 2020). Indeed, the World Health Organization reported that OA is the single most common cause of disability in the older population (Wittenauer, Smith and Aden 2013). The health burden of osteoarthritis will continue to rise with increasing obesity, sedentary lifestyle and an increasing growing population of 60 years or older, which is expected to double by 2050 (Wittenauer et al. 2013). It is a late-onset, complex disease of the joint that is typically characterised by degeneration and thinning of the articular cartilage, changes in synovium and subchondral bone causing loss in lubrication in a joint (Mandelbaum and Waddell 2005).

OA affects the whole joint but the main changes occur in articular cartilage leading to the complete loss of articular cartilage in severe cases. Articular cartilage is a connective tissue around 2 to 4 mm deep, found in joints of knees, hips, spine and fingers which plays the role of a shock absorber that facilitates transmission of loads and smooth painless movements of joints. Articular cartilage does not contain blood vessels, nerves, or lymphatics but is nourished by the synovial fluids, and is made up of a dense extracellular matrix that predominantly comprises water and electrolytes (60-85% by weight) as well as collagen, non-collagenous proteins, proteoglycans, lipids, phospholipids and a sparse distribution of highly specialized cells (chondrocytes, 10% by weight) that produce the matrix ((Bachrach et al. 1995, Mow, Ratcliffe and Poole 1992, Roughley 2001, Sophia Fox, Bedi and Rodeo 2009). The dry weight of the articular cartilage is primarily collagen (60%), however, the protein composition of cartilage is complex and varies depending on the depth of the tissue and distance from the chondrocytes (Quinn et al. 2013, Ellingsen et al. 2011, Müller et al. 2014).

Morphologically, cartilage can be divided into 3 layers: superficial, middle and deep (part of which is calcified, separated from the bone with a tidemark) zones and the key characteristics of each of these layers are summarized in Table 1 (Müller et al. 2014, Kato and Onodera 1988, Buckwalter and Mankin 1998, Hunziker 2002, Bhosale and Richardson 2008). While chondrocytes are found in all three layers, their shape, orientation and expression of collagen type as well as other matrix proteins vary depending on the layer of cartilage in which they are present (Goldring 2006, Müller et al. 2014, Sandell and Aigner 2001). Articular cartilage comprised of predominantly type II collagen (up to 90%) but it also contains types IX, XI and VI (Bielajew, Hu and Athanasiou 2020, Sandell and Aigner 2001, Eyre 2002). Type I and II collagens are fibrillar and have high activity in second harmonic generation (Chen et al. 2012).

**Table 1:** Collagen phenotype across cartilage layers.

| Cartilage Layer | % of the cartilage volume | Morphology of chondrocytes | Main collagen constituents |
| --- | --- | --- | --- |
| Superficial | 10-20% | Elongated/flattened | Mainly Type II (also, IX and XI) condensed collagen fibers running parallel to the joint |
| Middle | 40-60% | Round | Mainly Type II collagen fibers that are randomly arranged and slightly tilted. Transitioning from a more parallel (near the superficial zone) to a more perpendicular orientation (towards the deep layer). |
| Deep | ~30% | Round, arranged in columns | Type II collagen fibers running perpendicular to the joint and across the tidemark. |

Collagen type VI is found almost exclusively in the pericellular matrix (PCM), i.e., the area surrounding the chondron within which the chondrocytes are located providing structural integrity and facilitating communication with the extracellular matrix (Zhang 2015, Bielajew et al. 2020). PCM contains biglycan and 4 ecorin proteins which connect type VI collagen fibers with type II collagen fibers, providing stable matrix in the immediate proximity of chondrocytes (Zhang 2015).

At the onset of OA, two key changes are known to occur within the cartilage: (i) chondrocytes undergo proliferation, cell death, autophagy and form clusters (Goldring 2006, Sandell and Aigner 2001, Charlier et al. 2019) and, (ii) The fluid in the cavity of the joints, the synovial fluid, becomes rich in inflammatory cytokines, complement components and plasma proteins (Gobezie et al. 2007, Sohn et al. 2012). The primary inflammatory mediators have been identified as lipid molecules, prostaglandins and leukotriene (Laufer 2003, Wittenberg et al. 1993) and other molecules released by white adipose tissue (Pottie et al. 2006). Cartilage matrix degradation starts from the superficial layer and progresses to the deeper cartilage layers (Pritzker et al. 2006, Mankin et al. 1971, Buckwalter and Mankin 1998). However, the aetiology and molecular mechanisms responsible for the onset and progression of osteoarthritis remain poorly understood and their potential for diagnostic prediction has not been explored (Chen et al. 2017, Tong et al. 2022). The current gold-standard techniques for diagnosis of osteoarthritis focus on observation of the changes in morphology and bulk structure using advanced radiography, four-dimensional CT scan, CT arthrography, nuclear medicine techniques such as SPECT/CT, PET/CT, PET/MRI, three-dimensional quantitative cartilage morphometry; MRI and x-rays (Morris and Roessler 2006, Chen et al. 2017, Mandelbaum and Waddell 2005). These imaging techniques detect morphological changes such as narrowing of the space margin between the two bones in the joint as a consequence of cartilage loss or accumulation of synovial fluids and hence are typically limited to advanced stages of osteoarthritis, when the disease has progressed (Pritzker et al. 2006).

Early diagnosis of OA is highly desirable to enable timely implementation of lifestyle changes and medical interventions to reduce pain, improve mobility and patient quality of life. There is currently no cure for osteoarthritis and treatment regimens are targeted at alleviating inflammatory symptoms or arthroplasty such as a prosthetic joint replacement. Efficacious application of non-invasive and non-destructive techniques such as Raman spectroscopy that can directly (and potentially *in vivo*) detect subtle biochemical changes that occur within the cartilage before onset and during osteoarthritis and, critically will provide markers for diagnosis of osteoarthritis even before symptoms appear and thus aid the discovery of new pharmacological interventions to halt OA progression remains a key research goal. Raman spectroscopy (RS) is highly applicable for *in vivo* diagnosis as well as for evaluation of *ex vivo* articular cartilage samples due to its insensitivity to water and hence, can provide information about constituent molecules as well as their interactions with surrounding molecules despite its high water content. A number of characteristic Raman bands have been assigned to vibrational modes of constituent molecules in cartilage tissue. These include modes assigned to C-O stretching; amide I, random coil (1668 cm^-1^), Amide I, collagen secondary str (1640 cm^-1^), C=C stretching; phenylalanine, tryptophan (1606 cm^-1^), Amide II (1557 cm^-1^), CH2/CH3 scissoring; collagen and other proteins (1450 cm^-1^), COO^−^; GAGs (1424 cm^-1^), CH3; GAGs (1380 (proteoglycan) cm^-1^), (NH2) bending; amide III, α-helix (1270 cm^-1^), Amide III, α-helix (1260 cm^-1^), (NH2) bending; amide III, random coil (1245 cm^-1^), Amide III, random coil (1235 cm^-1^), Pyranose ring (1163 cm^-1^, 1042 cm^-1^), C-C, C-OH, C-N stretching, C-O-C glycosidic linkage (1125 cm^-1^), SO3^-^ stretching; GAGs (chondroitin sulfate) (1063 cm^-1^), Phenylalanine ring breathing (1003 cm^-1^), C-C stretching; collagen, α-helix (941 cm^-1^), C-C stretching; hydroxyproline (875 cm^-1^), C-C stretching; proline (858 cm^-1^) and C-C stretching; protein backbone (816 cm^-1^) (Nieuwoudt et al. 2020, Mansfield and Winlove 2017, Kumar et al. 2015, Pezzotti et al. 2022, Martinez et al. 2019, Khalid et al. 2018, Gao et al. 2021, Takahashi et al. 2014, Casal-Beiroa et al. 2021, Prokopi et al. 2021, Cárcamo et al. 2012, Asaoka et al. 2022).

For the diagnosis of osteoarthritis, it is important to identify the spectral changes in cartilage, that is, the changes in the peak patterns instead of intensity of individual peaks. This is carried out through multivariate analysis such as principal component and linear discriminant analysis (PCA and LDA) wherein the spectral loadings link the variance of the peaks across classes to the original spectra. Changes in the ratio of the intensity of certain vibrational modes also provide useful insight and can potentially provide diagnostic information. However, here we use PCA and LDA to understand the spectral differences between the different layers in healthy and OA cartilage for their unsupervised and supervised classification.

Raman Spectroscopy has been used by a number of groups to study human osteoarthritic cartilage (Takahashi et al. 2014, Gao et al. 2021, Prokopi et al. 2021, Bergholt et al. 2016, Casal- Beiroa et al. 2021, Kumar et al. 2015, Martinez et al. 2019). Takahashi et al completed a preliminary study on 5 arthritic human tibial cartilage samples retrieved from knee arthroplasty and found an increase in the relative intensity ratio between the Raman bands of collagen located at 1241 and 1269 cm^−1^ (amide III doublet) with increasing degradation grades indicating diagnostic potential (Takahashi et al. 2014). A recent feasibility study on 47 patient samples extended the aforementioned work and observed a decrease in sulfated glycosaminoglycans and proteoglycans and increase in collagen disorganization with severity of hip osteoarthritis (Casal-Beiroa et al. 2021). Similarly the diagnostic potential of Raman spectroscopy to study alterations in collagen structure in disease diagnosis was explored by multiple groups and reviewed by Martinez et al. (Martinez et al. 2019) but the different layers of cartilage were not taken into account. These studies confirm the diagnostic potential of Raman spectroscopy, however, to date, there has been no published evidence of the changes in the different layers of cartilage characterised by Raman spectroscopy or, the use of the differential signatures between the cartilage layers for osteoarthritis diagnosis in patients.

The current study has harnesses Raman Spectroscopy to characterize osteoarthritic and non-osteoarthritic patient samples of cartilage derived from femoral heads post hip arthroplasty. We investigated changes in the different layers of cartilage to identify discrete differences in the molecular composition. We used PCA and PC loadings to understand the contribution of different vibrational modes to unsupervised classification. We further carried out PCA-LDA for supervised classification. We examined the dependence of Raman spectral signatures from the different layers of cartilage with gender as well as age. We further used the multimodal imaging techniques of coherent anti-Stokes Raman scattering (CARS), second harmonic generation (SHG) and two-photon autofluorescence (TPF) microscopy on representative samples to correlate any changes in the structure and organization of different components in the superficial and deep layers with changes observed by Raman spectral analysis. The current work thus aims to explore the potential of OA diagnosis using the Raman signatures of different cartilage layers which can have significant diagnostic implications for an increasing aging demographic.

## 2. Experimental procedures

### 2.1. Sample preparation

Femoral heads were obtained with consent from patients undergoing total hip arthroplasty at Southampton General Hospital (SGH) and Spire Southampton Hospital, under Research Ethics Committee approval of the University of Southampton (18/NW/0231) and were clinically evaluated and classified as either osteoarthritic (Mankin score 3 to 4) or non-osteoarthritic.

All osteoarthritic donors (n=45, 24 female and 21 male) had no signs of osteoporosis or any other degenerative disease. All non-osteoarthritic donors (n=19, 10 female and 9 male) had osteoporosis but no obvious detectable osteoarthritis or other cartilage degenerative disease and, hence were treated as ‘healthy’ controls as confirmed by consultant orthopaedic surgeon (Professor Douglas Dunlop). The number and age of patients that donated samples for this study are shown in Table 1. Superficial and deep layer cartilage from non-weight bearing areas, were removed, under aseptic conditions using a scalpel blade (Fig.1), washed (PBS, 1X), fixed (4% formaldehyde,4° C, 72 h, gentle shaking), washed (PBS, 3X) and stored in PBS at 4°C in a refrigerator until Raman spectra were obtained.

**Figure 1.**
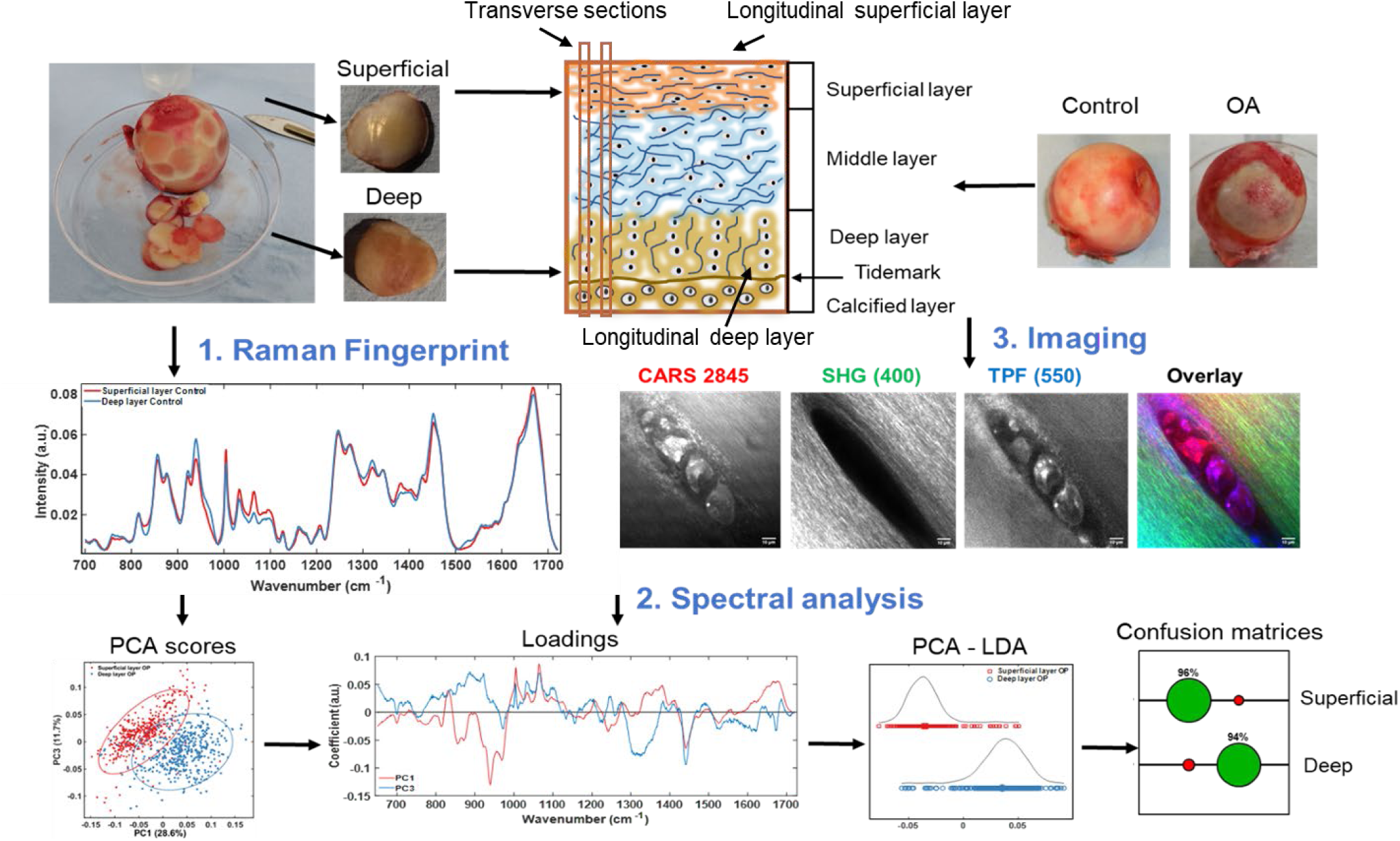
Workflow and methodology used in this work illustrated using OA cartilage samples. Top right shows femoral heads received post-surgery and the samples generated for analysis of superficial and deep layers (top left). A schematic shows the layered structure of the cartilage. (1). Shows the mean Raman spectra for the different cartilage layers (2). Spectra were analysed using Principal Component Analysis (PCA) and the scatter plot and loadings are shown. PCA was cascaded into Linear Discriminate Analysis (LDA) which improves the separation between classes. Confusion matrix shows accuracy of classification into superficial and deep layers based on its spectral signatures (3). Representative multimodal images of articular cartilage showing lipids and cell phospholipid bilayers (CARS), collagen fibres (SHG) and autofluorescent biological molecules (TPF). In overlay, the three modalities are combined: CARS (red), SHG (green), TPF (blue). Scale bar 10 μm.

**Table 1.**
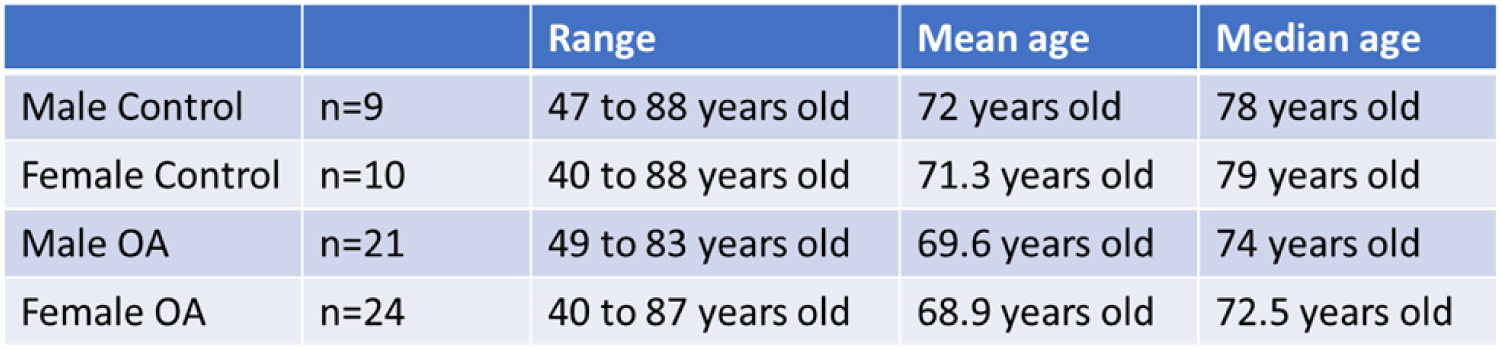
Age and gender of the patients whose samples were used in this study.

|  |  | Range | Mean age | Median age |
| --- | --- | --- | --- | --- |
| Male Control | n=9 | 47 to 88 years old | 72 years old | 78 years old |
| Female Control | n=10 | 40 to 88 years old | 71.3 years old | 79 years old |
| Male OA | n=21 | 49 to 83 years old | 69.6 years old | 74 years old |
| Female OA | n=24 | 40 to 87 years old | 68.9 years old | 72.5 years old |

### 2.2. Raman spectroscopy

Fixed cartilage slices (4 per patient) were chosen randomly, washed (distilled water, X2) and Raman spectra (fingerprint region including 615−1722 cm^−1^ and the CH2 stretching region from 2496−3265 cm^−1^) obtained (n= 15 to 20 per patient per superficial and deep layer) using a pre-calibrated (to 520 cm^−1^ reference peak of silicon) Renishaw InVia Raman microscope (785 nm laser, 10 mW, x50 (0.75 NA) short working distance (∼200 µm) objective, exposure 5 s, 3 accumulations) controlled by the proprietary Renishaw WIRE4.1 software, that enabled cosmic ray removal. Spectra were taken from the top (superficial layer) and then the sample was turned over for taking spectra from the bottom (deep layer) of the sample.

### 2.3. Principle Component Analysis

Principal Component Analysis (PCA) was performed using the IRootLab plugin (0.15.07.09-v) for MATLAB R2021b (Trevisan et al. 2013). All spectra were carefully background subtracted using a fifth-order polynomial, and the ends of each spectra were anchored to the axis using the rubber band function. Spectra were smoothed by wavelet denoising before vector normalization. Data was trained- mean-centred for PCA to provide a valid decomposition of the original data. PCA was then carried out with an optimal number of principle components (maximum set at 30). Percentages that give the portion of the data accounted for in a given PC (principal component or eigenvector) are displayed in figures and in Supplementary section. The PC loadings relate to the variation in the spectra to PC scores and thus indicate which regions in the Raman spectra make the highest contribution to PCA. In addition to the first principal component (PC1) that has the highest score and represents the maximum variance in the data, the PC with the next highest score was used to model the systematic variation of the data set (Lever, Krzywinski and Altman 2017).

### 2.4. Linear Discriminant Analysis

Linear Discriminant Analysis (LDA) was used to model differences within the RS data from superficial and deep layers of the cartilage samples following PCA analysis. Classes were assigned based on the PCA analysis. PCA is an unsupervised method that maximises the variation across the whole data set without taking into account the classes, if any, within a dataset. PCA-LDA cascade considers assigned classes and tries to maximize variation between them to give clustering and was used when PCA analysis alone failed to separate the data classes. While an unsupervised method is ideal for diagnosis, a supervised method such as PCA-LDA is likely to be closer to clinical application where a preliminary classification through clinical assessment or symptoms is likely to exist.

### 2.5. PCA Confusion matrices

Confusion matrices were created to predict the accuracy of the assignments of the Raman Spectra to a particular class/group based on the PCA scores. The analysis was undertaken following an online IRootlab tutorial “Classification with optimization of the number of “Principal Components” (PCs) aka PCA factors” (<u>PCA-LDC (studylib.net)</u> (Kaznowska et al. 2017). To account for the differences in the number of OA and Control sample numbers, Gaussian fit for unbalanced classes was applied followed by the k-fold cross validation. Optimal numbers of PCs were identified individually for each of the comparisons of the Raman spectra (e.g. Superficial vs Deep layer controls etc.) using the Random seed method and ranged from 10-30 PCs.

### 2.6. Multimodal imaging

Multimodal imaging was carried out to complement the findings of Raman spectroscopic analysis and to explore correlations to changes in architecture and arrangement of lipid structures (including cells) and collagen. Transverse sections of cartilage to the bone were taken with a scalpel blade and included superficial, middle and deep cartilage layers. The cartilage slices were washed with dH2O and positioned between two round cover slips (0.9 mm diameter, 0.17 thickness, Fisher) in a drop of dH2O fixed in a round ring sample holder (Attofluor™ Cell Chamber for microscopy Catalog number: A7816). Alternatively, cartilage slices were trimmed longitudinally with a razor blade to create flat were also taken from the top or the bottom. The samples were turned over on the sample stage for imaging and for Raman spectroscopy to acquire signals from top and bottom sequentially.

Images of cartilage slices were acquired using a home-built system which enabled image acquisition with coherent anti-Stokes Raman scattering (CARS), SHG and TPF, simultaneously. This multimodal laser scanning microscope employed ScanImage 5.1 (Vidrio Technologies) for image acquisition (Pologruto, Sabatini and Svoboda 2003). Briefly, for CARS imaging the fundamental of a fibre laser (1031 nm, 2 picosecond, 80 MHz, Emerald Engine, APE) was used as a Stokes beam, and the output of an optical parametric oscillator (OPO) (APE, Levante Emerald, 650–950 nm) which was synchronously pumped by the second harmonic (516 nm) of an AeroPulse fibre laser (NKT Photonics), was used as a pump beam. The multimodal platform utilises 2 ps pulses able to provide multimodal capability with CARS, SHG and TPF simultaneously. Non-linear interactions increase with shorter pulse-widths; however, with CARS since vibrational line-widths are of the order of 10 cm^-1^, 2 ps pulse-widths are near ideal to ensure efficient excitation and reduction of non-resonant background and at the same time limits any compromise of signal generation with other non-linear modalities such as SHG and TPF. The two beams were made collinear and then coupled through a galvanometric scanner to an inverted microscope (Nikon Ti-E) configured for epi-detection. Their temporal overlap was controlled with a delay line. For imaging lipids in the cartilage slices, the C–H stretching mode at 2845 cm^-1^ was targeted, and for this the OPO was tuned to 797.5 nm. The total incident power on the sample was approximately 60 mW. The SHG (400 nm) and TPF signal (550 nm) was collected with the same OPO beam at 797.5 nm simultaneously with CARS in a 3-channel detection setup equipped with photomultipliers (PMTs). Each sample was imaged using a x40 (1.15 NA) water immersion objective, with 3 or 6 optical zoom as specified in figures using galvanometric scanning, and acquisition time of 16 ms per line for a 512 x 512 pixel image. The detection setup included dichroic beamsplitters with cut-offs at 442 nm (Semrock Di02-R442) and 594 nm (Semrock Di02-R594).

## Results and Discussion

### 1. Methodology and workflow

Raman Spectroscopy (RS) was carried out to identify signatures of the superficial and deep layers of human cartilage from femoral heads. Cartilage from the femoral head of osteoporotic (OP) donor patients was used as a “healthy” control (Bielajew et al. 2020). Control femoral heads from OP patients, displayed a smooth uninterrupted cartilage surface and a fractured bone area at the “neck” of the sample (Fig. 1 (1)). Osteoarthritic (OA) femoral heads were predominantly Grade III-IV, with the cartilage completely absent from the weight bearing regions in most of the samples analysed (Fig. 1 (1). In order to investigate chemical changes prior to complete disappearance of cartilage only intact cartilage was removed and used for Raman spectroscopy. All analysis was carried out on age and gender matched samples to the extent possible given the distribution of patients undergoing hip arthroplasty (Table 1). The mean age for all the groups was around 70 years old. Median age for control (OP) samples was around 5 years higher than for OA samples, it ranged from 72.5 years for female OA and 74 years old for male OA; while it was 78 and 79 years old for male and female controls respectively (Table 1).

The workflow involved: (i) the acquisition of Raman Spectra and multimodal images from the superficial and deep layers, (ii) analysis of spectral data (between 700 cm^-1^ to 1720 cm^-1^) using principal component and linear component analysis and classification (Fig.1) (iii) identification of peaks/bans that account for differences from the PCA loadings and determination of classification accuracy (iv) correlating spectral analysis with multi-modal images using CARS/SHG/TPF. PCA (unsupervised) and scatter plots using PC1 and PC2 (or the principal component that captures the most variation) scores helped to classify the data, while PCA-LDA scatter plot showed clear separation of the classes (i.e., superficial vs deep, healthy vs OA, female vs male and over 60’s vs under 60’s) (Fig.1(2)). Confusion matrices were created from PCA where the size of the circles indicates classifications (green) and misclassifications (red), respectively. Cross-sections of representative cartilage samples were imaged using CARS/SHG/TPF multimodal imaging to observe any chemical and structural changes that can support Raman spectral analysis. CARS was used to image structures rich in lipids by using the symmetric stretch vibration of –CH2 bonds at 2845 cm^-1^; SHG was used to image collagen fibres and TPF was used to image autofluorescence emitted at 550 nm that highlights cells rich in FAD/NADH (Qin and Xia 2021, Schaefer et al. 2019). Representative images are shown in Fig.1(3) wherein collagen fibres can be seen around a cluster of 4 cells with different levels of lipids and autofluorophores.

### 2. Raman signatures of superficial and deep layer in healthy cartilage

The methodology described above was applied to superficial and deep cartilage layers of each slice in healthy cartilage samples (Fig.2 A). The mean spectra from the superficial and deep layers were compared to identify differences between their “Raman fingerprint”. A prominent difference between superficial and deep layers at a sulphated glycosaminoglycan (sGAG) peaks of 1064 cm^-1^ and 1380 cm^-1^ was observed which are indicative of different mechanical and biochemical matrix properties as shown in Klein et al. and others (Klein et al. 2007, Bachrach et al. 1995, Gao et al. 2021, Sharma et al. 2007). The concentration of aggrecan, which is a large proteoglycan and contains large chondroitin sulphate chains (1064 cm^-1^ peak) is much higher in the superficial layer to accommodate extracellular water retention capacity to counteract external pressures of the loads onto the cartilage (Roughley and Mort 2014). Seven other peaks were also identified (asterisks Fig. 2B and in Loadings in Fig. 2E), corresponding to 877 cm^-1^ (Hydroxyproline), 921 cm^-1^ (Proline), 938 cm^-1^ (Hydroxyproline), 1004 cm^-1^ (Phenylalanine), 1320 cm^-1^ (Collagen wagging)/1345 cm^-1^ (Collagen twisting), 1451 cm^-1^ (Collagen type II/Amide III), 1666 cm^-1^ (Collagen/Amide I). These Raman bands were the main contributors to PC1 and PC3 scores in Fig. 2C as evident from Loadings in Fig. 2E, except for the 1004 cm^-1^ Phenylalanine peak which displayed variability within spectral readings. LDA analysis clearly delineated superficial and deep layer spectra on the LD1 axis with negligible overlap (Fig. 2D). The confusion matrix based on PCA showed an accuracy of 96% and 94% for superficial and deep layers, respectively.

**Figure 2.**
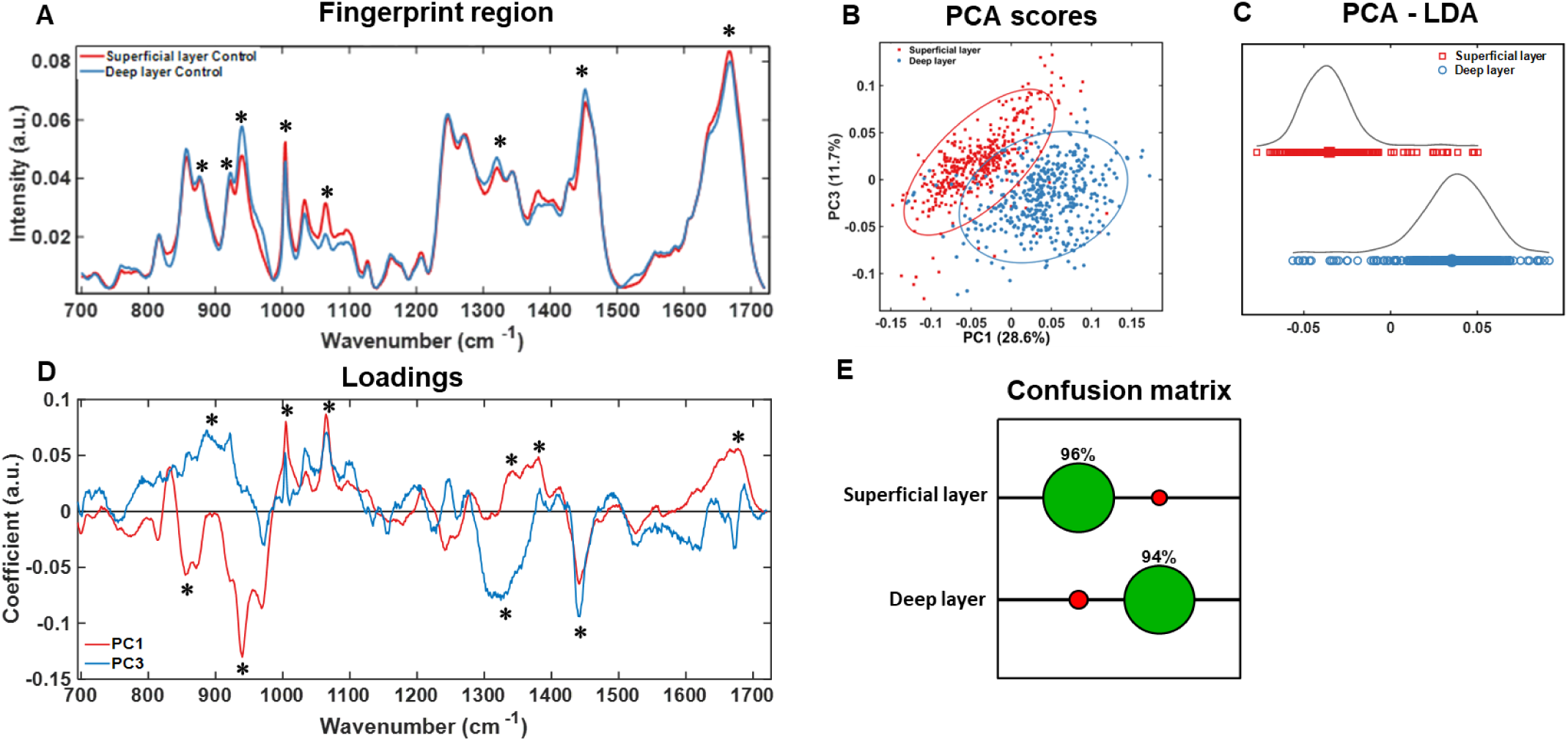
Raman spectroscopic analysis of superficial and deep layers of healthy cartilage. (A) Fingerprint region (700 to 1720 cm^-1^) of RS of superficial (red) and deep (blue) layers of healthy “control” cartilage. (B) 2-D PCA scatter plot shows distribution of superficial and deep layer spectra along PC1 and PC3 axes. (C) LDA analysis shows further separation of superficial (red) and deep (blue) layers based on class labels. (D) PC1 and PC3 loadings from the PCA. (E) Confusion matrix shows the classification accuracy with correct classifications indicated in green and mis- classifications indicated in red of spectra from superficial or deep layer cartilage. N=19 (male n=9, female n=10). Asterisks (“*”) in (A) refer to “*” in (D) to highlight the main spectral peaks that contribute to PCA in (B).

### 3. Raman signatures of superficial and deep layer in OA cartilage

OA cartilage samples were analysed similar to healthy samples to find out if there were any differences in Raman signatures between the different layers. The spectra and analysis results are shown in Figure 3. Prominent differences between superficial and deep layers were found at the sulphated glycosaminoglycan (sGAG) peaks of 1064 cm^-1^ and 1380 cm^-1^ and hydroxyproline peak of 938 cm^-1^, with the latter peak potentially representing a change in collagen structure in OA. Six other peaks (indicated with asterisks in Fig. 3A and D) were found to make high contributions to PC1 and PC2 loadings; the peak positions and likely assignments are as follows: 855 cm^-1^ (Proline/Collagen backbone), 877 cm^-1^ (Hydroxyproline), 921 cm^-1^ (Proline), 938 cm^-1^ (Hydroxyproline), 1320 cm^-1^ (Collagen wagging)/1345 cm^-1^ (Collagen twisting), 1451 cm^-1^ (Collagen type II/Amide III), 1666 cm^-1^ (Collagen/Amide I). LDA analysis clearly separated the superficial and deep layer spectra along the LD1 axes (Fig. 3C). Confusion matrix on PCA showed an accuracy of 91% for the classification of both the superficial and deep layers.

**Figure 3.**
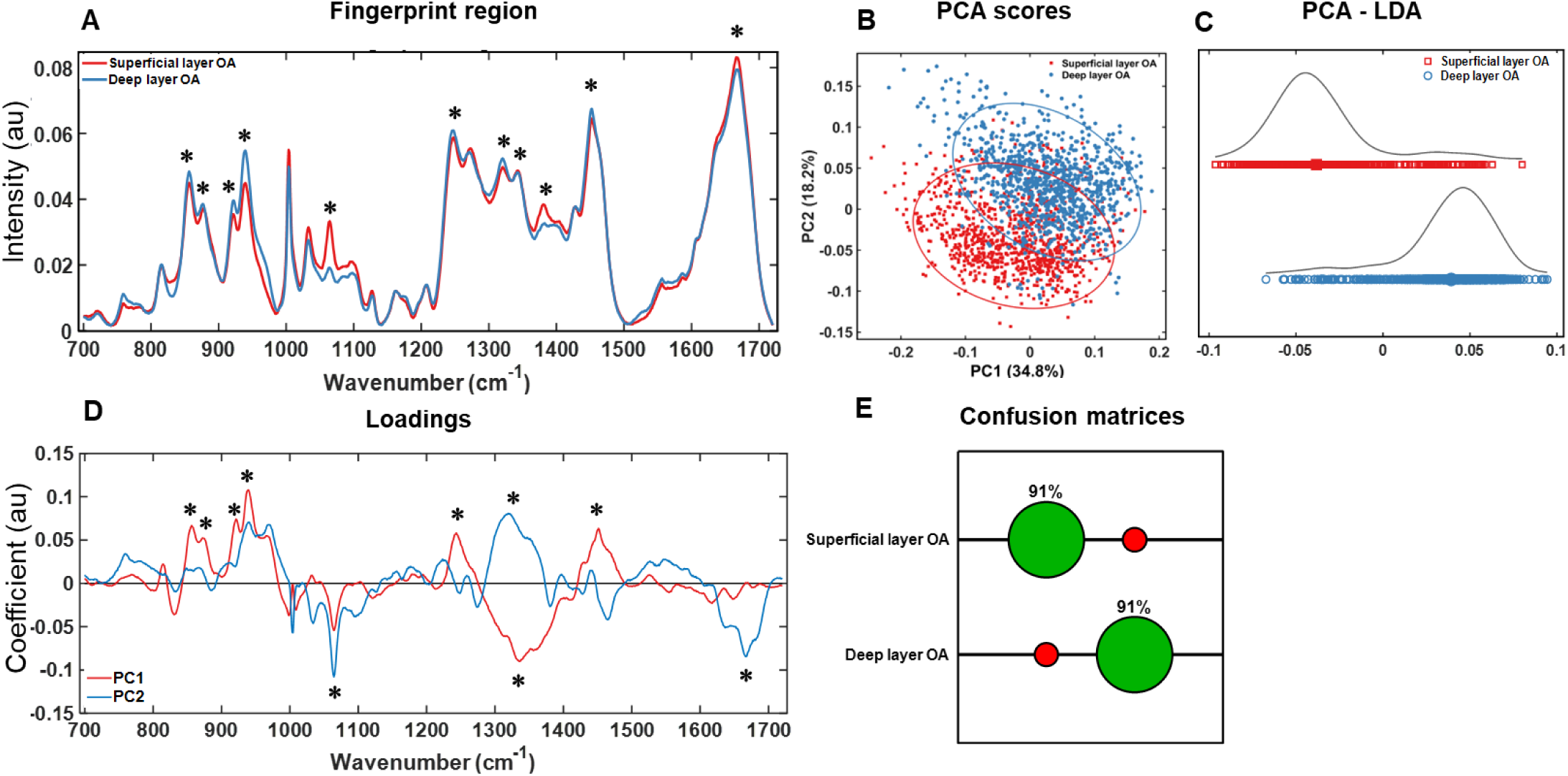
Raman spectroscopic analysis of superficial and deep layers of osteoarthritic (OA) cartilage. (A) Fingerprint region (700 to 1720 cm^-1^) of RS of superficial (red) and deep (blue) layers of OA cartilage. (B) 2-D PCA scatter plot showing the distribution of superficial and deep layer spectra along PC1 and PC2 axes. (C) LDA analysis showed clear separation of superficial (red) and deep (blue) layer spectra into the respective classes. (D) PC1 and PC2 loadings corresponding to PCA scatter plot shown in (C). (E) Confusion matrix shows correct classifications (green) and mis- classifications (red) of superficial and deep layer spectra from OA cartilage samples. N=45 (male n=21, female n=24). Asterisks (“*”) in (A) refer to “*” in (D) and point to spectral peaks contributing to PCA scores in (B).

### 4. Diagnostic signatures of OA identified from Raman spectroscopy of superficial and deep layers of cartilage

To determine the diagnostic potential of Raman signatures at different depths, we compared and analysed the spectra from OA and control superficial and deep layers of cartilage (Fig. 4). Results indicated that the main peaks that differentiated superficial and deep cartilage layers were 1320 cm^-1^ (Collagen wagging) and 1345 cm^-1^ (Collagen twisting) peaks of collagen modes. This is consistent with remodelling of “fibrillar structures” collagen in OA (Kumar et al. 2015, Takahashi et al. 2014) and characterised by initiation of synthesis of type IIa and III procollagens that are less efficient (Sandell and Aigner 2001). In addition, another 6 peaks were observed to be key in distinguishing OA from healthy cartilage in the Raman Spectra of the superficial layer, specifically 921 cm^-1^ (Proline), 938 cm^- 1^ (Hydroxyproline), 1004 cm^-1^ (Phenylalanine), 1380 cm^-1^ and 1064 cm^-1^ (sulphated glycosaminoglycan (sGAG), 1451 cm^-1^ (Collagen type II/Amide III). Proline, hydroxyproline, phenylalanine and sulphated glycosaminoglycan are known to undergo changes in OA (Asaoka et al. 2022, Casal-Beiroa et al. 2021, Kumar et al. 2015, Gao et al. 2021, Pezzotti et al. 2022). Changes in sGAG are indicative of osmolarity/hydration changes that occur in cartilage with disease (Robin Poole et al. 1989, Wong et al. 1996, Kumar et al. 2015, Roughley 2001, Roughley and Mort 2014), while amide III peaks relate to amounts of total collagen in the samples. 1004 cm^-1^ phenylalanine and the 1245 cm^-1^ collagen peaks have been reported to be altered (Gaifulina et al.). We found less significant differences between them compared to sulphated glycosaminoglycan (sGAG) 1064 cm^-1^ peak. Depth correlated comparisons carried out in this study have potentially led to the different results and insights. It is clear from the spectral loadings (Suppl. Fig. 1A) that sGAG (1380 cm^-1^ peak) is increased in OA superficial layer compared to controls. It has also been assigned to sGAGs (Asaoka et al. 2022) and indicates an unstable cartilage matrix in OA. (Fig. 4 C, Loadings in suppl. Fig. 1 (1A). Additionally,1272 cm^-1^ peak Loadings were altered in both superficial and deep layers consistent with the remodelling/change of direction of collagen (Unal, Jung and Akkus 2015, Pezzotti et al. 2022, Takahashi et al. 2014) (Fig. 4C, Loadings in suppl. Fig. 1 (1A, 2A). In summary, changes in collagen and matrix (sGAG) peaks in the superficial layer and changes mainly of the collagen peaks constitute a diagnostic signature for OA. Further analysis by PCA-LDA allows classification of healthy cartilage with an accuracy of 80% from either the analysis of superficial or deep layers while OA cartilage was identified with an increased accuracy of 84% from the analysis of superficial and 87% from deep layer.

**Figure 4.**
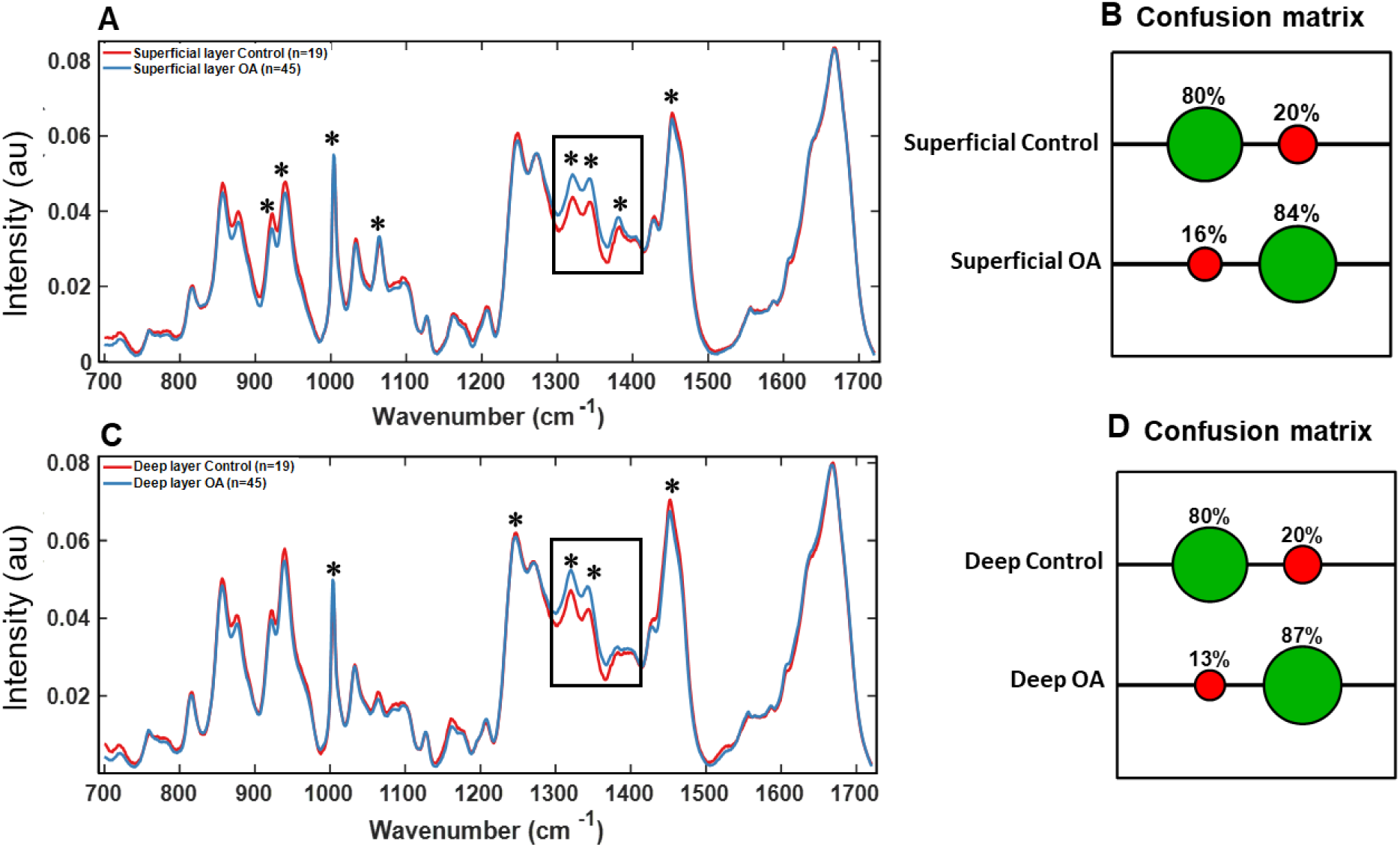
Raman fingerprint for OA diagnosis of articular cartilage. (A) RS of superficial layer and (C) of deep layer from Control (red) vs OA (blue) cartilage. Boxed regions show the region with the highest difference for OA and contains 1320/1340 collagen modes and 1380 sGAG (C) identified by asterisks. Confusion matrixes in (B) and (D) show classification accuracy with correct assignments indicated in green and mis-assignments indicated in red. Additional “*” in (A) and in (C) refer to loadings in Supplementary Figure S1 labelled with “*” and indicate spectral peaks that contribute to PCA scores (Suppl.1 and 2 B).

### 5. Effect of gender on Raman fingerprint of different cartilage layers

To establish that the OA diagnostic signatures were not related to gender-specific differences Raman spectra from gender matched healthy cartilage samples were compared. The mean Raman spectra from the superficial and deep layers in healthy cartilage are shown in Fig. 5A and C. The main peaks 921 cm^-1^ (Proline), 938 cm^-1^ (Hydroxyproline), 1004 cm^-1^ (Phenylalanine), 1064 cm^-1^ (sulphated glycosaminoglycan (sGAG), 1320 cm^-1^ (Collagen wagging) and 1340 cm^-1^ (Collagen twisting) modes, 1666 cm^-1^ (Collagen/Amide I) that contributed to PCA loadings (Supplementary Fig. S2). Importantly, these peaks also distinguish control and OA samples as shown in Fig. 4, except for the 1666 cm^-1^ peak that is indicative of total protein content in cartilage. However, the Raman spectroscopy data and the PCA analysis, demonstrated fewer differences in chemical composition between genders.

**Figure 5.**
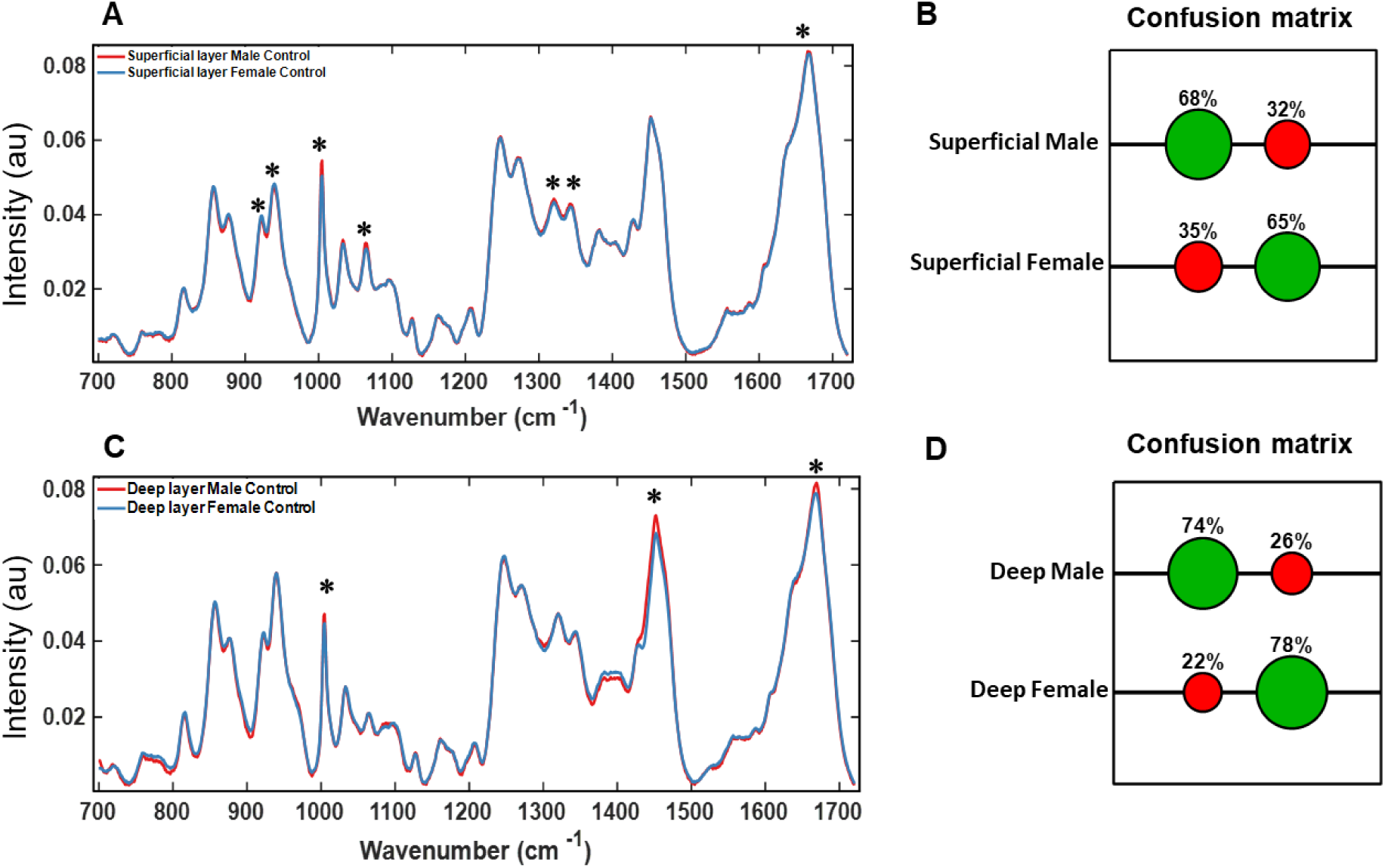
Effect of gender on Raman signatures in different layers of healthy articular cartilage. (A) RS of superficial layer and (C) of deep layers from healthy cartilage samples of male (red) vs female (blue). Confusion matrixes in (B) and (D) demonstrate positive (green circles) and negative (red circles) scores in assignments to Male or Female samples. Asterisks (“*”) in (A) refer to 1(A) and in (C) to 2(A) Loadings in Supplementary figure 2 labelled with “*” and show spectral peaks that contribute to PCA scores (Suppl.2, 1A and 2A).

The peaks correspond to protein composition and structure of collagen fibres, which may be more sensitive to changes occurring in OA. Interestingly, proline, sGAG and collagen structural modes contributed to PCA loadings for the superficial layer but not for the deep layer. The large overlap in PCA scores (Fig S2A) in the male and female samples indicated a high level of similarity between the spectra. Consequently the accuracy of classification was modest, at 74% and 78% for the deep layer and 68% and 65% for the superficial layer (Fig 5B and D). Since there is a larger overlap and lesser accuracy of classification with gender it is indicative that OA results in significant changes in chemical composition across both genders resulting in lesser overlap in PCA and higher accuracy of classification as shown in the previous section (Fig 4).

### 6. Raman spectroscopy identifies age-related chemical fingerprint in healthy and OA

Age is an important factor in manifestation of OA. Hence, we also examined whether there were any age-related differences in the different layers of cartilage in healthy and OA cartilage. Samples were divided into two age groups (under 60 and over 60 years). The groups were selected to provide a cohort aged below 60 years of age (“young”) and an “older” cohort, over 60 years of age given the prevalence of OA increases in older patients (Fig 6).

**Figure 6.**
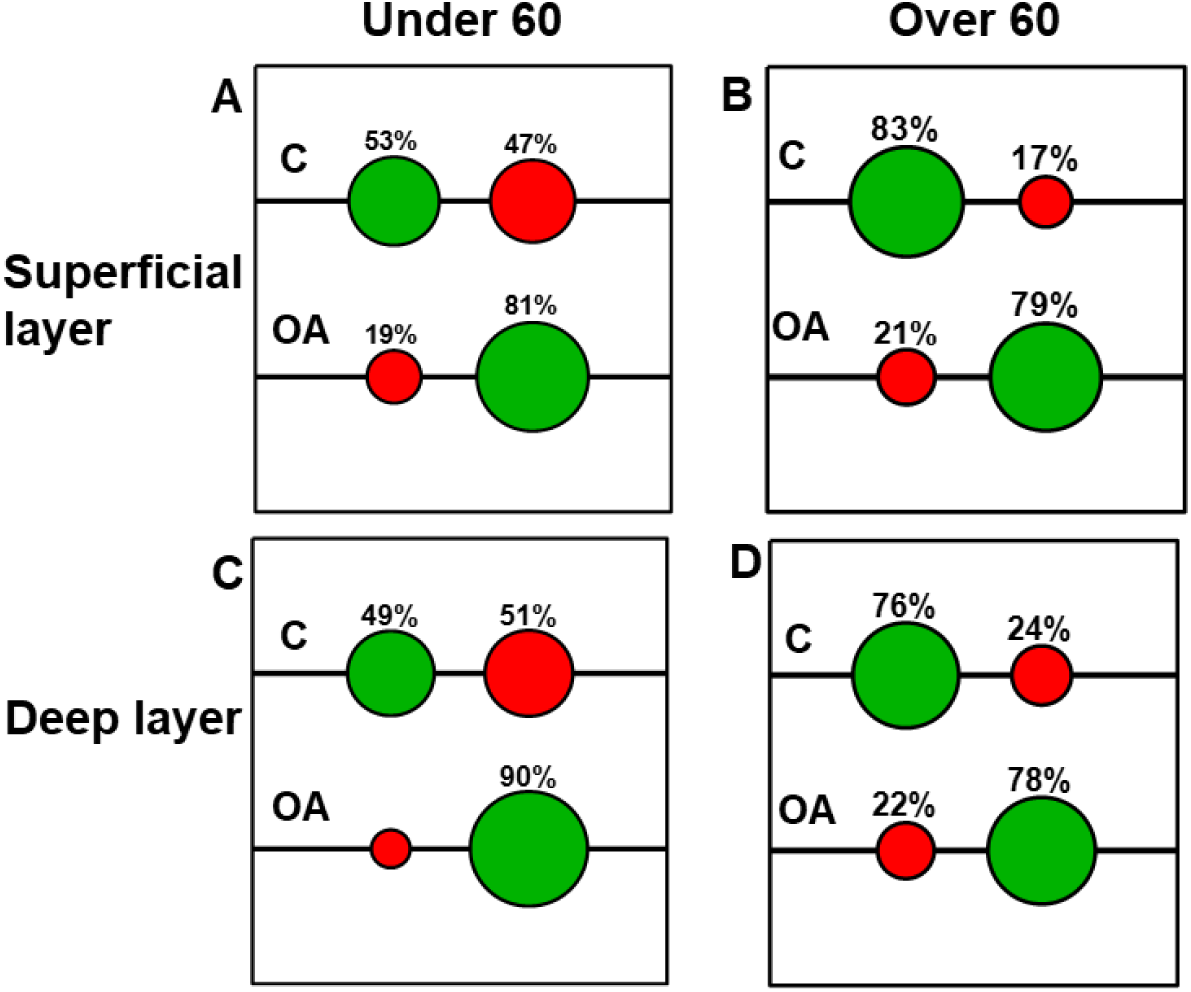
Effect of age on Raman spectral signatures of different cartilage layers. Confusion matrices based on Raman spectra for OA diagnosis in under 60 and over 60 patient age groups. Superficial layer (A) and Deep layer (C) in under 60 cohort (control n=5, OA n=13). Superficial layer (B) and Deep layer (D) in over 60 cohort (control n=14, OA n=32). Confusion matrices demonstrate percentage accuracy of classification (green) and mis-classifications (red) based on PCA of RS. C – control (healthy cartilage) samples.

The under 60 patient cohort, included 5 healthy and 13 OA samples and the over 60 patient cohort comprised of 14 healthy and 32 OA samples. Spectra from under 60 OA samples were correctly classified with 90% accuracy from superficial and 81% accuracy from deep layer analysis while spectra from over 60 OA patient samples were correctly assigned with 79% accuracy from superficial and 78% accuracy from deep layer analysis. On the other hand, spectra from under 60 healthy samples were correctly assigned with 53% accuracy from superficial and 49% accuracy from the deep layer while that from over 60 healthy samples were correctly assigned with 83% accuracy from superficial and 76% accuracy from deep layer analysis. PCA and LDA analysis is shown in Fig. S3 and S4. The classification accuracy was low for spectra from under 60 healthy samples potentially due to lower sample numbers (n=5). This analysis shows that spectra from the superficial layer of over 60 healthy patient samples while spectra from the deep layer of under 60 OA patient samples had more diagnostic information.

### 7. Multimodal imaging of cross sections of cartilage from Superficial to deep layer

To investigate the differences in observed in the spectra further, CARS (2845 cm^-1^), SHG (400 nm) and Two photon fluorescence (550 nm) images of transverse section (section perpendicular to the cartilage/bone interface) and in longitudinal direction from both the top (superficial layer) and bottom (deep layer) of the sample (as shown in Fig. 1) were acquired (3 controls and 4 OA patients) for a preliminary analysis (Figure 7 and 8).

**Figure 7.**
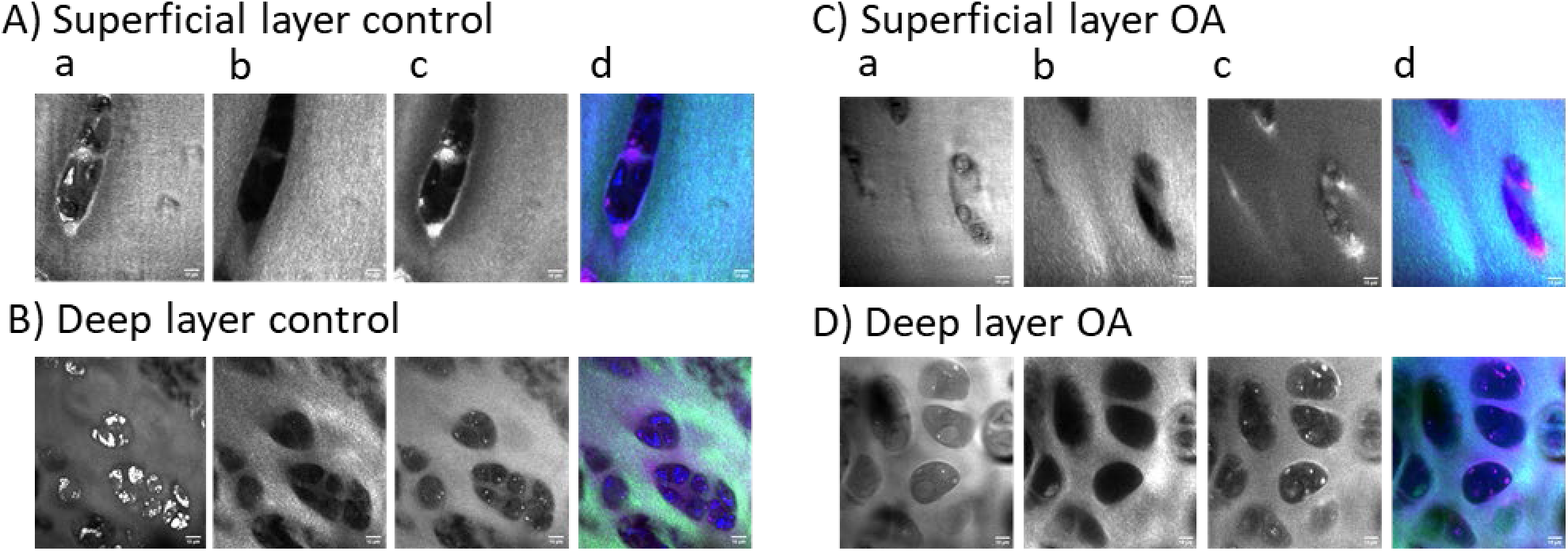
Multimodal imaging of transverse (perpendicular) sections of cartilage. A) Superficial and B) Deep layer for a control patient sample (M84 OP); C) Superficial and D) Deep layer for an OA patient sample (F44 OA). (a) CARS at 2845 cm^-1^ showing lipids, cell membranes and matrix, (b) SHG (400 nm) showing type II collagen fibres in cartilage, (c) TPH (550 nm) showing autofluorescence of collagen and other biological molecules, (d) overlay with CARS (red), SHG (green) and TPF (blue). The scale bar is 10 μm.

**Figure 8.**
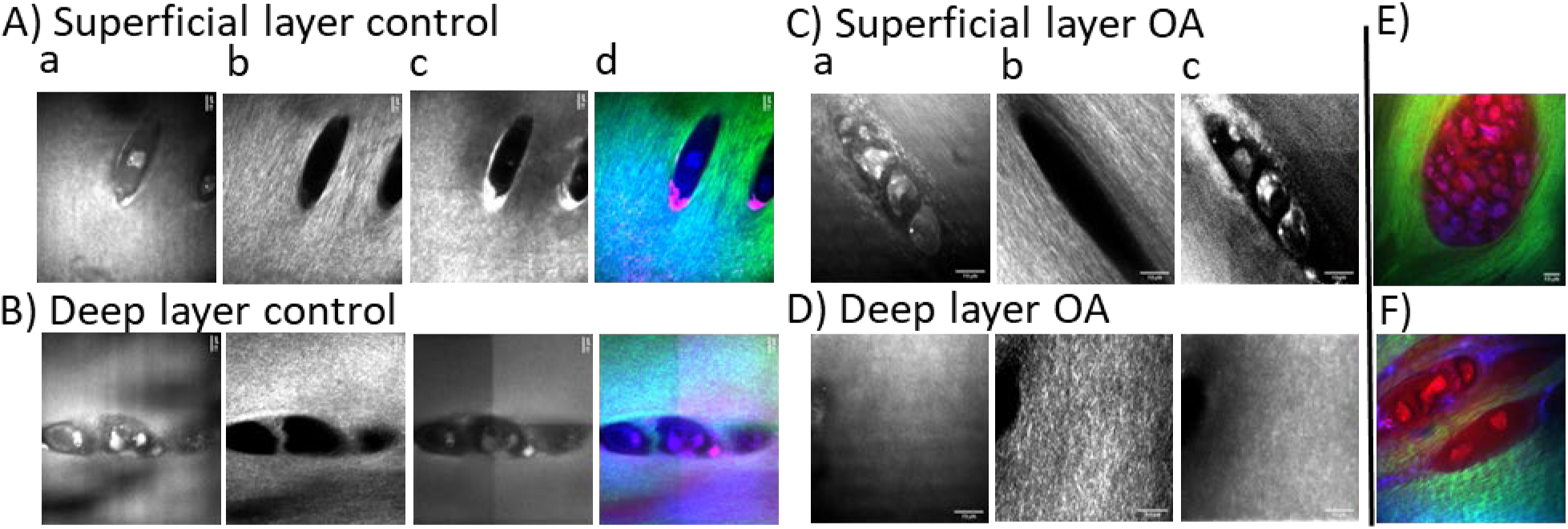
Multimodal imaging in the longitudinal direction (from the top and the bottom) of cartilage. A) Superficial and B) Deep layer for a control sample (M88 OP); C) Superficial and D) Deep layer for a OA sample (F69 OA). (a) CARS at 2845 cm^-1^ showing lipids, cell membranes and cartilage matrix, (b) SHG (400 nm) showing type II collagen fibres, (c) TPH (550 nm) showing autofluorescence of collagen and other biological molecules, (d) overlay with CARS (red) of cells, SHG (green) of collagen and TPF (blue) of autofluorescent biological molecules. E) and F) OA example from M63 and M64 patients showing clustered cells (red) and ‘wavy’ collagen fibres (green). The scale bar is 10 μm.

In the transverse sections (Figure 7), the collagen fibres in the SHG images appeared as discrete white streaks (potentially bundles of tightly packed fibres) within a grey/white background and notably more abundant in superficial compared to deep cartilage layers. Outlines of cells were distinguishable in CARS (lipid membranes) and TPF (autofluorescence from pericellular components such as collagen VI and/or NADH/FADH) images. Lipid droplets inside the cells were present in both controls and OA samples. In figure 7B) (a) bright lipid droplets within chondrocytes in the deep layer of M84 control cartilage were not observed in any other samples and potentially represents a patient specific result unrelated to any other conditions. We noted the shape and cluster appearance of the chondrocytes in the superficial (large, horizontal and elongated) and deep (small, clustered, vertical) layers differed in direction and size and were in agreement with published research on stained cartilage (Sandell and Aigner 2001). Similarly, representative images of the top (superficial layer) and bottom (deep layer) of cartilage in healthy and OA cartilage samples are shown in Figure 8. CARS and TPF images were comparable to that shown in Figure 7, however, SHG images (panel b) showed more defined and increased brightness of collagen fibres in longitudinal compared to transverse direction. This effect is due to the direction of the collagen fibres and their alignment with the section direction. The superficial layer likely contains fibres that form a meshwork and hence, can be seen in both the longitudinal and transverse directions. However, the collagen fibres are aligned along the longitudinal direction in the deep layer of cartilage and hence, appear diffuse in the transverse section while showing their expected fibre-like structures only in the images taken longitudinally. In the SHG images of the deep layer, the collagen fibres appeared more ‘wavy’ in OA than in controls patient samples. 7E) and 7F) are examples from different OA patients and illustrate cell clustering, a signature of OA (Sandell and Aigner 2001). The “wavy” appearance of collagen fibres may offer a new diagnostic feature for OA. Such imaging-based signatures will be explored with additional control and OA samples. Nevertheless, the current observations with the representative multimodal imaging data contain clear changes in collagen in both superficial and deep layers across control and OA samples and indicate correlation with the Raman spectroscopic analysis.

### 8. Conclusions

In this work we have used Raman Spectroscopy and multi-modal imaging (CARS, SHG and TPF) to characterize osteoarthritic and non-osteoarthritic human cartilage derived from femoral heads post hip arthroplasty and have investigated changes in the superficial and deep layers We have identified discrete differences in the signatures that are likely due to changes in collagen and other matrix biological molecules such as sGAGs. Specifically, the vibrational peaks/modes corresponding to 877 (hydroxyproline), 1064 cm^-1^ and 1380 cm^-1^ (sGAG), 921 cm^-1^ (proline, prominent in collagen) and 1245, 1320, 1345, 1451, 1666 cm^-1^ (collagen) account for the differences between the different cartilage layers as well as between OA and healthy cartilage. Raman spectroscopic measurements and multivariate analysis indicated clear differences between the chemical composition of superficial and deep layers of cartilage with an accuracy of >94% classification. Comparison of superficial layer between healthy and OA samples indicated a >84% accuracy in classification achieved for OA. The spectral differences between the superficial and deep layers showed less differences and reduced accuracy with gender but clear differences between below 60 and above 60 age groups. The relatively larger overlap and lesser accuracy of classification based on gender shown in healthy cartilage samples indicates that larger chemical changes across both genders occur due to OA in different cartilage layers. Age based analysis indicated the importance of analysing different layers; superficial layer analysis resulted in higher accuracy of classification for healthy samples in over 60 while deep layer analysis resulted in higher accuracy of classification for under 60. Multimodal imaging techniques confirmed changes in the structure and organization of the chondrocytes and collagen in the superficial and deep layers that correlated with the results from Raman spectral analysis. In summary, the current work establishes the potential of OA diagnosis using the Raman signatures of different cartilage layers. Our work also shows that chemical changes in different cartilage layers is informative could allow early diagnosis with label-free technique such as Raman spectroscopy.

## Supporting information

Supplementary Information including figures

## Data Availability

All data produced in the present study are available upon reasonable request to the authors

## References

Asaoka, R., H. Kiyomatsu, H. Miura, A. Jono, T. Kinoshita, M. Takao, T. Katagiri & Y. Oshima (2022) Prognostic potential and pathological validation of a diagnostic application using Raman spectroscopy in the characterization of degenerative changes in the cartilage of the humeral head. J Biomed Opt, 27.

Bachrach, N. M., W. B. Valhmu, E. Stazzone, A. Ratcliffe, W. M. Lai & V. C. Mow (1995) Changes in proteoglycan synthesis of chondrocytes in articular cartilage are associated with the time-dependent changes in their mechanical environment. Journal of Biomechanics, 28, 1561–1569.

Bergholt, M. S., J.-P. St-Pierre, G. S. Offeddu, P. A. Parmar, M. B. Albro, J. L. Puetzer, M. L. Oyen & M. M. Stevens (2016) Raman Spectroscopy Reveals New Insights into the Zonal Organization of Native and Tissue-Engineered Articular Cartilage. ACS Central Science, 2, 885–895.

Bhosale, A. M. & J. B. Richardson (2008) Articular cartilage: structure, injuries and review of management. Br Med Bull, 87, 77–95.

Bielajew, B. J., J. C. Hu & K. A. Athanasiou (2020) Collagen: quantification, biomechanics and role of minor subtypes in cartilage. Nature Reviews Materials, 5, 730–747.

Buckwalter, J. A. & H. J. Mankin (1998) Articular cartilage: degeneration and osteoarthritis, repair, regeneration, and transplantation. Instr Course Lect, 47, 487–504.

Cárcamo, J. J., A. E. Aliaga, R. E. Clavijo, M. R. Brañes & M. M. Campos-Vallette (2012) Raman study of the shockwave effect on collagens. Spectrochim Acta A Mol Biomol Spectrosc, 86, 360–5.

Casal-Beiroa, P., V. Balboa-Barreiro, N. Oreiro, S. Pértega-Díaz, F. J. Blanco & J. Magalhães (2021) Optical Biomarkers for the Diagnosis of Osteoarthritis through Raman Spectroscopy: Radiological and Biochemical Validation Using Ex Vivo Human Cartilage Samples. Diagnostics, 11, 546.

Charlier, E., C. Deroyer, F. Ciregia, O. Malaise, S. Neuville, Z. Plener, M. Malaise & D. de Seny (2019) Chondrocyte dedifferentiation and osteoarthritis (OA). Biochem Pharmacol, 165, 49–65.

Chen, D., J. Shen, W. Zhao, T. Wang, L. Han, J. L. Hamilton & H.-J. Im (2017) Osteoarthritis: toward a comprehensive understanding of pathological mechanism. Bone Research, 5, 16044.

Chen, X., O. Nadiarynkh, S. Plotnikov & P. J. Campagnola (2012) Second harmonic generation microscopy for quantitative analysis of collagen fibrillar structure. Nature Protocols, 7, 654–669.

Ellingsen, P. G., M. B. Lilledahl, L. M. Aas, L. Davies Cde & M. Kildemo (2011) Quantitative characterization of articular cartilage using Mueller matrix imaging and multiphoton microscopy. J Biomed Opt, 16, 116002.

Eyre, D. (2002) Collagen of articular cartilage. Arthritis Res, 4, 30–5.

Gao, T., A. J. Boys, C. Zhao, K. Chan, L. A. Estroff & L. J. Bonassar (2021) Non-Destructive Spatial Mapping of Glycosaminoglycan Loss in Native and Degraded Articular Cartilage Using Confocal Raman Microspectroscopy. Frontiers in Bioengineering and Biotechnology, 9.

Gobezie, R., A. Kho, B. Krastins, D. A. Sarracino, T. S. Thornhill, M. Chase, P. J. Millett & D. M. Lee (2007) High abundance synovial fluid proteome: distinct profiles in health and osteoarthritis. Arthritis Res Ther, 9, R36.

Goldring, M. B. (2006) Update on the biology of the chondrocyte and new approaches to treating cartilage diseases. Best Pract Res Clin Rheumatol, 20, 1003–25.

Hunter, D. J., L. March & M. Chew (2020) Osteoarthritis in 2020 and beyond: a Lancet Commission. The Lancet, 396, 1711–1712.

Hunziker, E. (2002) Articular cartilage repair: basic science and clinical progress. A review of the current status and prospects osteoarthritis and cartilage, 10.

Kato, M. & T. Onodera (1988) Morphological investigation of osteochondrosis induced by ofloxacin in rats. Fundamental and Applied Toxicology, 11, 120–131.

Kaznowska, E., J. Depciuch, K. Szmuc & J. Cebulski (2017) Use of FTIR spectroscopy and PCA-LDC analysis to identify cancerous lesions within the human colon. Journal of Pharmaceutical and Biomedical Analysis, 134, 259–268.

Khalid, M., T. Bora, A. A. Ghaithi, S. Thukral & J. Dutta (2018) Raman Spectroscopy detects changes in Bone Mineral Quality and Collagen Cross-linkage in Staphylococcus Infected Human Bone. Scientific Reports, 8, 9417.

Klein, T. J., M. Chaudhry, W. C. Bae & R. L. Sah (2007) Depth-dependent biomechanical and biochemical properties of fetal, newborn, and tissue-engineered articular cartilage. Journal of Biomechanics, 40, 182–190.

Kumar, R., K. M. Grønhaug, N. K. Afseth, V. Isaksen, C. de Lange Davies, J. O. Drogset & M. B. Lilledahl (2015) Optical investigation of osteoarthritic human cartilage (ICRS grade) by confocal Raman spectroscopy: a pilot study. Analytical and Bioanalytical Chemistry, 407, 8067–8077.

Laufer, S. (2003) Role of eicosanoids in structural degradation in osteoarthritis. Curr Opin Rheumatol, 15, 623–7.

Lever, J., M. Krzywinski & N. Altman (2017) Principal component analysis. Nature Methods, 14, 641–642.

Mandelbaum, B. & D. Waddell (2005) Etiology and pathophysiology of osteoarthritis. Orthopedics, 28, s207–14.

Mankin, H. J., H. Dorfman, L. Lippiello & A. Zarins (1971) Biochemical and Metabolic Abnormalities in Articular Cartilage from Osteo-Arthritic Human Hips: II. CORRELATION OF MORPHOLOGY WITH BIOCHEMICAL AND METABOLIC DATA. JBJS, 53.

Mansfield, J. C. & C. P. Winlove (2017) Lipid distribution, composition and uptake in bovine articular cartilage studied using Raman micro-spectrometry and confocal microscopy. Journal of Anatomy, 231, 156–166.

Martinez, M. G., A. J. Bullock, S. MacNeil & I. U. Rehman (2019) Characterisation of structural changes in collagen with Raman spectroscopy. Applied Spectroscopy Reviews, 54, 509–542.

Morris, M. D. & B. J. Roessler (2006) Future spectroscopic diagnostics in osteoarthritis. Future Rheumatology, 1, 383+.

Mow, V. C., A. Ratcliffe & A. R. Poole (1992) Cartilage and diarthrodial joints as paradigms for hierarchical materials and structures. Biomaterials, 13, 67–97.

Müller, C., A. Khabut, A. Aspberg, D. Heinegård, J. Dudhia & P. Önnerfjord (2014) Quantitative proteomics of different zones in human articular cartilage reveals unique patterns of protein distribution. Osteoarthritis and Cartilage, 20, S36.

Nieuwoudt, M. K., R. Shahlori, D. Naot, R. Patel, H. Holtkamp, C. Aguergaray, M. Watson, D. Musson, C. Brown, N. Dalbeth, J. Cornish & M. C. Simpson (2020) Raman spectroscopy reveals age- and sex-related differences in cortical bone from people with osteoarthritis. Scientific Reports, 10, 19443.

Pezzotti, G., W. Zhu, Y. Terai, E. Marin, F. Boschetto, K. Kawamoto & K. Itaka (2022) Raman spectroscopic insight into osteoarthritic cartilage regeneration by mRNA therapeutics encoding cartilage-anabolic transcription factor Runx1. Materials Today Bio, 13, 100210.

Pologruto, T. A., B. L. Sabatini & K. Svoboda (2003) ScanImage: flexible software for operating laser scanning microscopes. Biomed Eng Online, 2, 13.

Pottie, P., N. Presle, B. Terlain, P. Netter, D. Mainard & F. Berenbaum (2006) Obesity and osteoarthritis: more complex than predicted! Ann Rheum Dis, 65, 1403–5.

Pritzker, K. P. H., S. Gay, S. A. Jimenez, K. Ostergaard, J. P. Pelletier, P. A. Revell, D. Salter & W. B. van den Berg (2006) Osteoarthritis cartilage histopathology: grading and staging. Osteoarthritis and Cartilage, 14, 13–29.

Prokopi, N., K. S. Andrikopoulos, A. S. Beobide, G. A. Voyiatzis & D. J. Papachristou (2021) Collagen orientation probed by polarized Raman spectra can serve as differential diagnosis indicator between different grades of meniscus degeneration. Scientific Reports, 11, 20299.

Qin, Y. & Y. Xia (2021) Simultaneous Two-Photon Fluorescence Microscopy of NADH and FAD Using Pixel-to-Pixel Wavelength-Switching. Frontiers in Physics, 9.

Quinn, T. M., H. J. Häuselmann, N. Shintani & E. B. Hunziker (2013) Cell and matrix morphology in articular cartilage from adult human knee and ankle joints suggests depth-associated adaptations to biomechanical and anatomical roles. Osteoarthritis Cartilage, 21, 1904–12.

Robin Poole, A., Y. Matsui, A. Hinek & E. Lee (1989) Cartilage macromolecules and the calcification of cartilage matrix. The anatomical record, 224, 167–179.

Roughley, P. J. (2001) Articular cartilage and changes in arthritis: noncollagenous proteins and proteoglycans in the extracellular matrix of cartilage. Arthritis Res, 3, 342–7.

Roughley, P. J. & J. S. Mort (2014) The role of aggrecan in normal and osteoarthritic cartilage. Journal of Experimental Orthopaedics, 1, 8.

Sandell, L. J. & T. Aigner (2001) Articular cartilage and changes in Arthritis: Cell biology of osteoarthritis. Arthritis Research & Therapy, 3, 107.

Schaefer, P. M., S. Kalinina, A. Rueck, C. A. F. von Arnim & B. von Einem (2019) NADH Autofluorescence—A Marker on its Way to Boost Bioenergetic Research. Cytometry Part A, 95, 34–46.

Sharma, A., L. D. Wood, J. B. Richardson, S. Roberts & N. J. Kuiper (2007) Glycosaminoglycan profiles of repair tissue formed following autologous chondrocyte implantation differ from control cartilage. Arthritis Research & Therapy, 9, R79.

Sohn, D. H., J. Sokolove, O. Sharpe, J. C. Erhart, P. E. Chandra, L. J. Lahey, T. M. Lindstrom, I. Hwang, K. A. Boyer, T. P. Andriacchi & W. H. Robinson (2012) Plasma proteins present in osteoarthritic synovial fluid can stimulate cytokine production via Toll-like receptor 4. Arthritis Res Ther, 14, R7.

Sophia Fox, A. J., A. Bedi & S. A. Rodeo (2009) The basic science of articular cartilage: structure, composition, and function. Sports Health, 1, 461–8.

Takahashi, Y., N. Sugano, M. Takao, T. Sakai, T. Nishii & G. Pezzotti (2014) Raman spectroscopy investigation of load-assisted microstructural alterations in human knee cartilage: Preliminary study into diagnostic potential for osteoarthritis. J Mech Behav Biomed Mater, 31, 77–85.

Tong, L., H. Yu, X. Huang, J. Shen, G. Xiao, L. Chen, H. Wang, L. Xing & D. Chen (2022) Current understanding of osteoarthritis pathogenesis and relevant new approaches. Bone Research, 10, 60.

Trevisan, J., P. P. Angelov, A. D. Scott, P. L. Carmichael & F. L. Martin (2013) IRootLab: a free and open-source MATLAB toolbox for vibrational biospectroscopy data analysis. Bioinformatics, 29, 1095–1097.

Unal, M., H. Jung & O. Akkus (2015) Novel Raman Spectroscopic Biomarkers Indicate that Post-Yield Damage Denatures Bone’s Collagen. Journal of Bone and Mineral Research, 31.

Wittenauer, R., L. Smith & K. M. Aden. 2013. Background Paper 6.12 Osteoarthritis.

Wittenberg, R. H., R. E. Willburger, K. S. Kleemeyer & B. A. Peskar (1993) In vitro release of prostaglandins and leukotrienes from synovial tissue, cartilage, and bone in degenerative joint diseases. Arthritis Rheum, 36, 1444–50.

Wong, M., P. Wuethrich, P. Eggli & E. Hunziker (1996) Zone-specific cell biosynthetic activity in mature bovine articular cartilage: A new method using confocal microscopic stereology and quantitative autoradiography. Journal of Orthopaedic Research, 14, 424–432.

Zhang, Z. (2015) Chondrons and the pericellular matrix of chondrocytes. Tissue Eng Part B Rev, 21, 267–77.

