## Supplementary Information including figures for "Harnessing Raman spectroscopy and Multimodal Imaging of Cartilage for Osteoarthritis Diagnosis"

Anna Crisford[1,2], Hiroki Cook[1], Konstantinos Bourdakos[1], Seshasailam Venkateswaran[1], Douglas Dunlop[2], Richard OC Oreffo*[2, 3] Sumeet Mahajan*[1, 3].
[1]School of Chemistry, Faculty of Engineering and Physical Sciences; [2]Human Development and Health, Faculty of Medicine; [3]Institute for Life Sciences, University of Southampton, United Kingdom

### Supplementary Information

Table 1. Summary of classification accuracies of Raman spectra.

| **Cartilage sample** | **Healthy (Control)** | **OA** |
| --- | --- | --- |
| **Superficial layer in Healthy (Control)** | **96%** | **94%** |
| **Deep layer in Healthy (Control)** | **91%** | **91%** |
| **Superficial layer in Healthy (Control) compared to OA** | **80%** | **84%** |
| **Deep layer in Control compared to OA** | **80%** | **87%** |
| **Superficial layer in under 60** | **53%** | **81%** |
| **Deep layer in under 60** | **49%** | **90%** |
| **Superficial layer in over 60** | **83%** | **79%** |
| **Deep layer in OA over 60** | **76%** | **78%** |
| **Superficial layer in Male** | **68%** | **na** |
| **Superficial layer in Female** | **65%** | **na** |
| **Deep layer in Male** | **74%** | **na** |
| **Deep layer in Female** | **78%** | **na** |

Table 2. Main peak assignments in Raman spectra of superficial and deep layers in human articular cartilage

| **Frequency ($cm^{-1}$)** | **Assignments to chemical bonds and molecules** |
|---|---|
| 855 | -C-C- Proline ring (P) |
| 877 | -C-C- Hydroxyproline ring (HP) |
| 921 | -C-C- Proline ring (P) |
| 938 | -C-C- Hydroxyproline ring (HP) |
| 1004<br>1046 | Phenylalanine ring breathing mode of collagen and proteoglycans |
| 1033 | Phenylalanine ring breathing mode of collagen and proteoglycans, differences in collagen content |
| 1064 | $SO_3$ stretching, Chondroitin sulphate (CH) (part of sGAG complex)<br>-C-C- skeletal saturated fatty acid |
| 1085 | -C-C- skeletal unsaturated fatty acid |
| 1101 | Hyaluronic acid (HA)<br>/Chondroitin sulphate (CS) /Heparan sulfate (HS) |
| 1128 | HA<br>-C-C- skeletal saturated fatty acid |
| 1245 | CN stretching of amide bond, Amide III, random coil (disordered) |
| 1272 | NH deformation of amide bond, Amide III α-helix or coil (ordered) |
| 1320 | $CH_2$, $CH_3$ twisting, Amide III, collagen twisting mode |
| 1345 | $CH_2$ scissoring, Amide III, collagen bending mode. GAGS (Glycosaminoglycans) |
| 1380 | Lipid<br>GAGS (Glycosaminoglycans) |

| | |
|---|---|
| 1451 | $CH_2$, $CH_3$ scissoring, $NH_2$ deformation of amide bond, collagen and other proteins |
| 1666 | $NH_2$ deformation, Amide I (Collagen) |

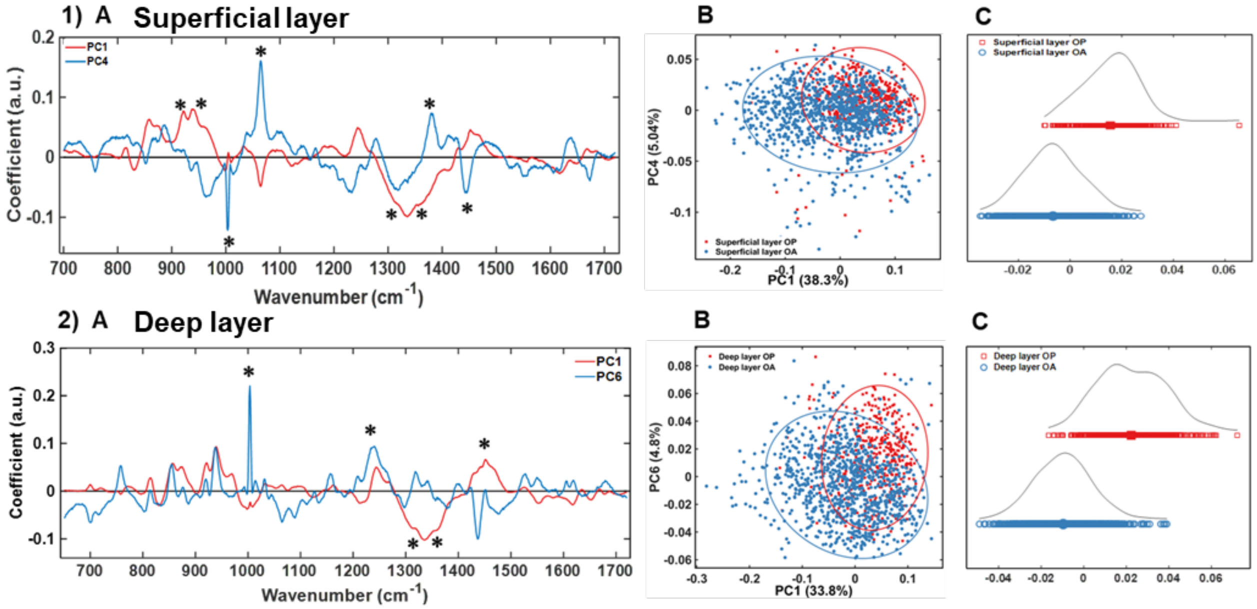


**Supplementary figure 1 (to Figure 4).** Raman fingerprint for OA diagnosis of articular cartilage. 1) Superficial and 2) Deep layer (A) Loadings to demonstrate main peaks contributing to PCA of RS spectral points in 1)B and 2)B. 1)C superficial and 2)C deep layer LDA analysis separates spectra into positive and negative quadrants based on classes labels from PCA analysis in B). OA (blue) vs Control (red) cartilage. Peaks with the highest variability are framed. Confusion matrixes in (B) and (D) demonstrate Positive and negative scores in layers assignments.

"*" in (A) show spectral peaks contribution to PCA scores (B).

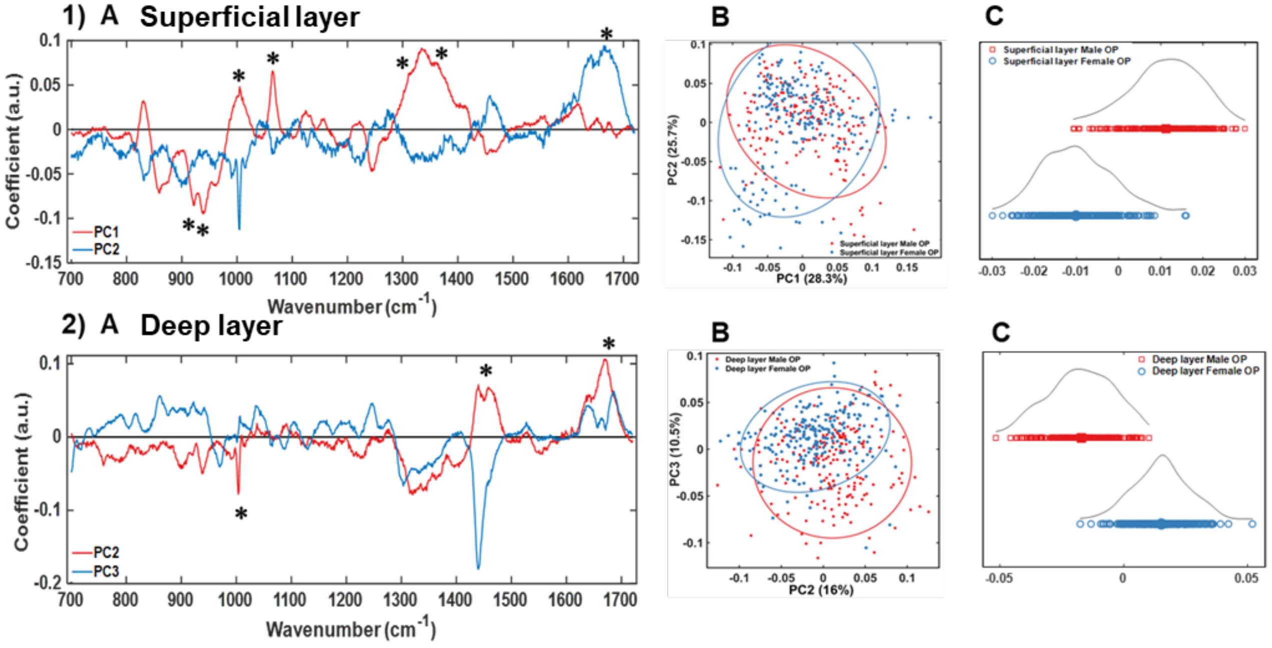


**Supplementary figure 2 (to Figure 5).** Raman fingerprint for articular cartilage from gender related samples. 1(A) superficial and 2(A) deep layer Loadings to demonstrate main peaks contributing to PCA of RS spectral points in 1)B and 2)B. 1)C superficial and 2)C deep layer LDA analysis separates spectra into positive and negative quadrants based on classes labels from PCA analysis in B). Cartilage samples from Male (red) vs Female (blue) samples. "*" in (A) show spectral peaks contribution to PCA scores (B).

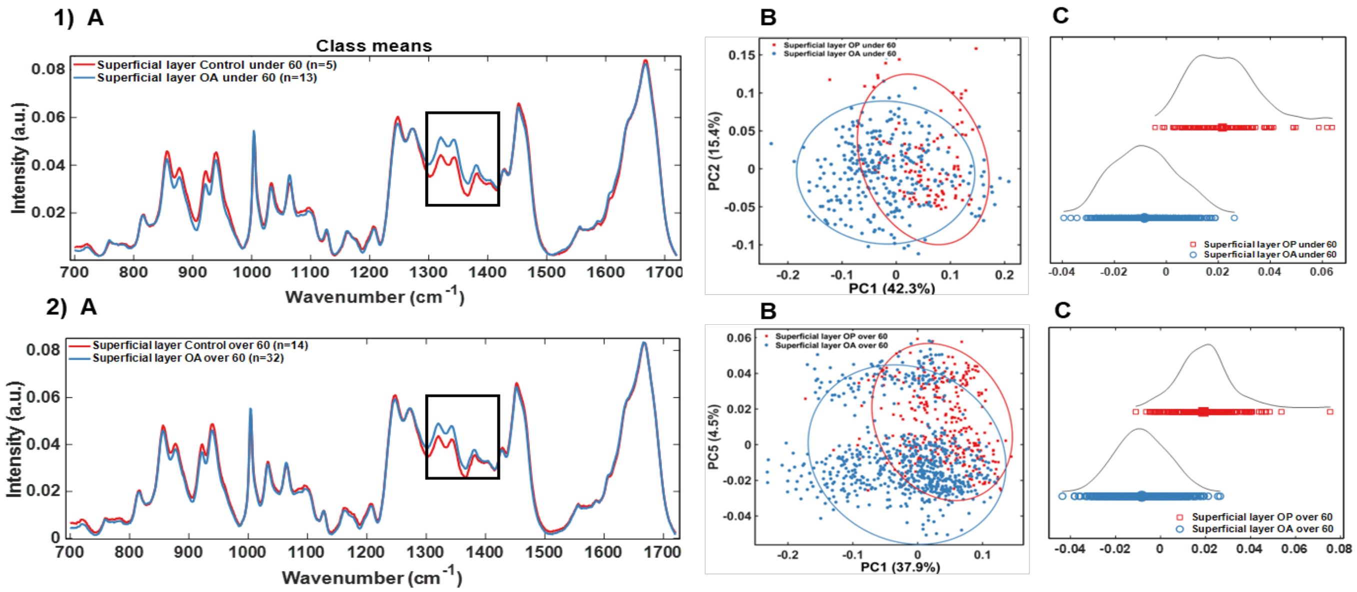


**Supplementary figure 3 (to Figure 6).** Raman fingerprint from superficial layer of articular cartilage from age matched samples. RS of 1(A) superficial layer, from under 60 years old (control n=5, OA n=13) and 2(A) superficial layer, from over 60 years old (control n=14, OA n=32) individuals. Boxed spectra (1320$cm^{-1}$, 1345$cm^{-1}$, 1380$cm^{-1}$ peaks) show highest differences between control and OA samples. 2(A) and 2(B) 2-D representation of PCA analysis of RS spectral points from 1)A and 2)A. LDA 1-D representation of 1(C) superficial layer, from under 60 years old and 2(C) superficial layer, from over 60 years old individuals separates spectra into positive and negative quadrants based on classes labels from PCA analysis in B). Cartilage samples from Controls 1(A) superficial layer, from under 60 years old and 2(A) superficial layer, from over 60 years old individuals. Male (red) vs Female (blue) samples.

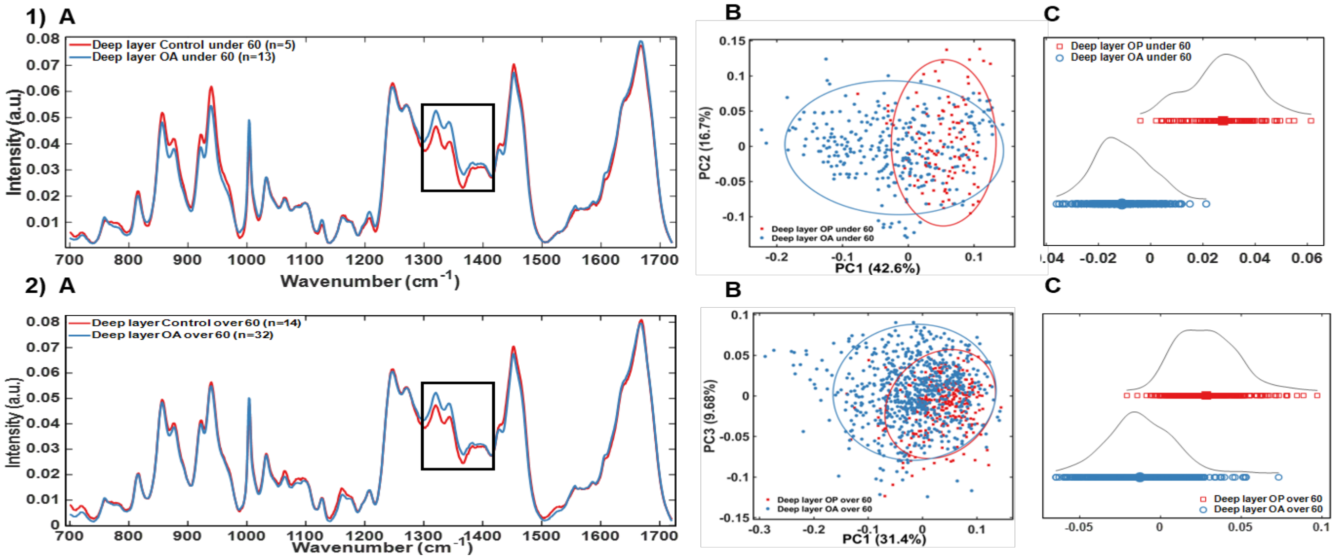


**Supplementary figure 4 (to Figure 6).** Raman fingerprint from deep layer of articular cartilage from age matched samples. RS of 1(A) superficial layer, from under 60 years old (control n=5, OA n=13) and 2(A) superficial layer, from over 60 years old (control n=14, OA n=32) individuals. Boxed spectra (1320$cm^{-1}$, 1345$cm^{-1}$, 1380$cm^{-1}$ peaks) show highest differences between control and OA samples. 2(A) and 2(B) 2-D representation of PCA analysis of RS spectral points from 1)A and 2)A. LDA 1-D representation of 1(C) superficial layer, from under 60 years old and 2(C) superficial layer, from over 60 years old individuals separates spectra into positive and negative quadrants based on classes labels from PCA analysis in B). Cartilage samples from Controls 1(A) superficial layer, from under 60 years old and 2(A) superficial layer, from over 60 years old individuals. Male (red) vs Female (blue) samples.

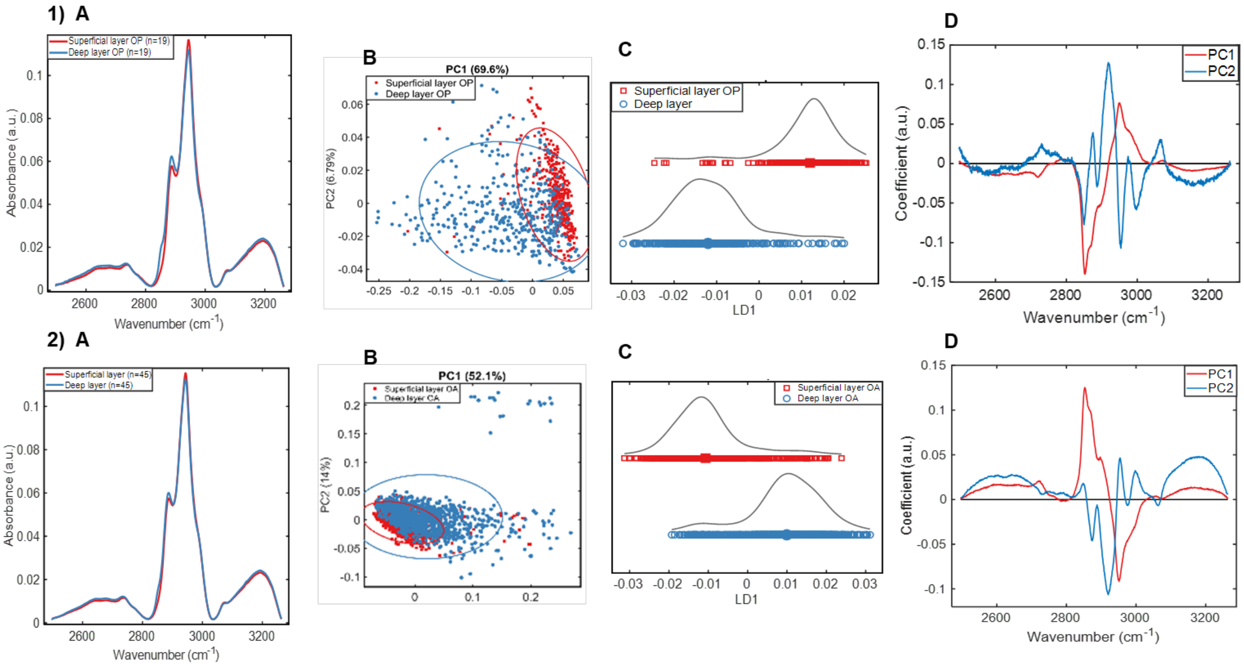


**Supplementary figure 5.** Raman spectra at CH region for deep tissue imaging of articular cartilage. Top panel (1) control samples (n=19), bottom panel (2) OA samples (n=45). 1, 2 (A) RS of CH region (2500 to 3300) of superficial (red) and deep (blue) layers of cartilage. 1, 2 (B) 2-D scatter plot demonstrating distribution of superficial and deep layer spectra along PC1 and PC2 axes. 1, 2 (C) LDA analysis shows further separation of superficial (red) and deep (blue) layers based on class labels from PCA. (D) Loadings from PC1 and PC2 PCA scores in B). Control samples: mean age 72 years, median age 78 years, n=19 (male=9, female=10). OA samples: mean age 69 years, median 73 years, n=45 (male=21, female=24).

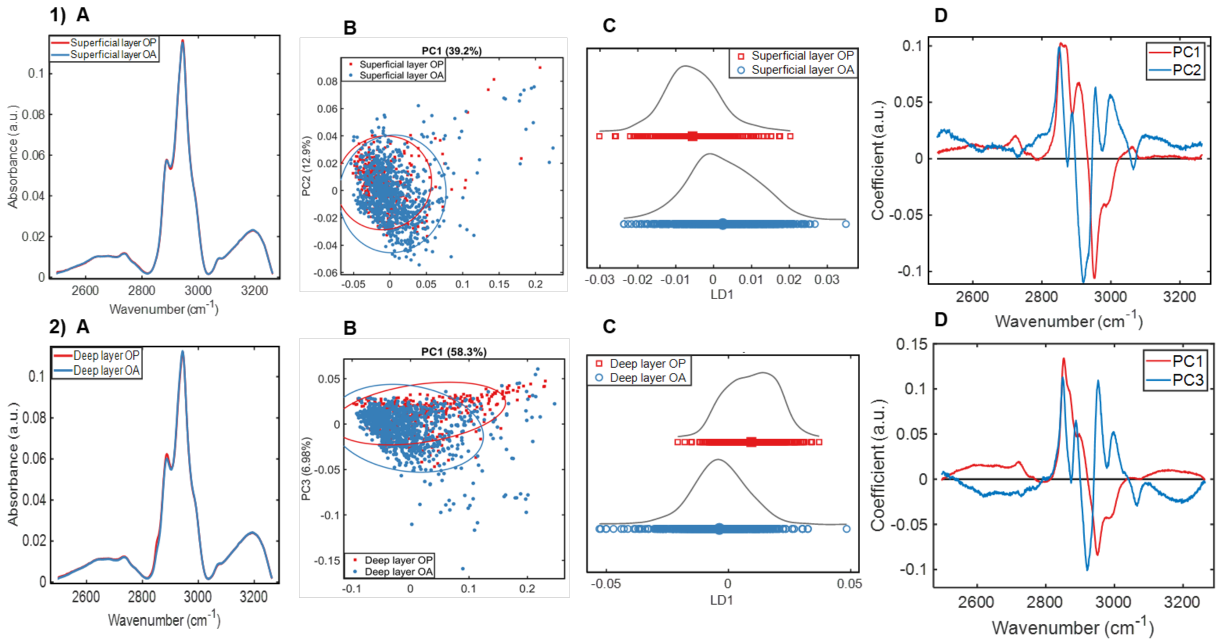


**Supplementary figure 6.** Raman spectra at CH region for OA diagnosis of articular cartilage. 1, 2 (A) RS of CH region (2500 to 3300) of control (red, n=19) vs OA (blue, n=45) samples; superficial layer in top panel (1), deep layer in bottom panel (2). 1, 2 (B) 2-D scatter plot demonstrating distribution of superficial and deep layer spectra along PC1/PC2 in (1B) and PC1/PC3 in (2B) axes. 1, 2 (C) 1-D LDA analysis of superficial (1C) and deep (2C) control (red) and deep (blue) layers based on class labels. 1(D) Loadings from PC1 and PC2; 2(D) from PC1 and PC3 PCA scores in (B). Control samples: mean age 72 years, median age 78 years, n=19 (male=9, female=10). OA samples: mean age 69 years, median 73 years, n=45 (male=21, female=24).
